# Genome-wide Association Study identifies two novel loci for Gilles de la Tourette Syndrome

**DOI:** 10.1101/2021.12.11.21267560

**Authors:** Fotis Tsetsos, Apostolia Topaloudi, Pritesh Jain, Zhiyu Yang, Dongmei Yu, Petros Kolovos, Zeynep Tumer, Renata Rizzo, Andreas Hartmann, Christel Depienne, Yulia Worbe, Kirsten R. Müller-Vahl, Danielle C. Cath, Dorret I. Boomsma, Tomasz Wolanczyk, Cezary Zekanowski, Csaba Barta, Zsofia Nemoda, Zsanett Tarnok, Shanmukha S. Padmanabhuni, Joseph D. Buxbaum, Dorothy Grice, Jeffrey Glennon, Hreinn Stefansson, Bastian Hengerer, Evangelia Yannaki, John A. Stamatoyannopoulos, Noa Benaroya-Milshtein, Francesco Cardona, Tammy Hedderly, Isobel Heyman, Chaim Huyser, Pablo Mir, Astrid Morer, Norbert Mueller, Alexander Münchau, Kerstin J. Plessen, Cesare Porcelli, Veit Roessner, Susanne Walitza, Anette Schrag, Davide Martino, The TSAICG, The TSGeneSEE initiative, The EMTICS collaborative group, The TS-EUROTRAIN network, The TIC Genetics collaborative group, Jay A. Tischfield, Gary A. Heiman, A. Jeremy Willsey, Andrea Dietrich, Lea K. Davis, James Crowley, Carol A. Mathews, Jeremiah M. Scharf, Marianthi Georgitsi, Pieter J. Hoekstra, Peristera Paschou

**Affiliations:** Department of Molecular Biology and Genetics, Democritus University of Thrace, Alexandroupolis, Greece; Department of Biological Sciences, Purdue University, West Lafayette, IN, USA; Psychiatric and Neurodevelopmental Genetics Unit, Center for Genomic Medicine, Department of Psychiatry, Massachusetts General Hospital, Boston, MA, USA; Stanley Center for Psychiatric Research, Broad Institute of MIT and Harvard, Cambridge, MA, USA; Department of Clinical Genetics, Kennedy Center, Copenhagen University Hospital, Rigshospitalet, Denmark; Department of Clinical Medicine, Faculty of Health and Medical Sciences, University of Copenhagen; Child and Adolescent Neurology and Psychiatry, Department of Clinical and Experimental Medicine, University of Catania, Catania, Italy; Department of Neurology, Hôpital de la Pitié-Salpêtrière, Paris, France; Institute for Human Genetics, University Hospital Essen, Essen, Germany; Assistance Publique Hôpitaux de Paris, Hopital Saint Antoine, Paris France; French Reference Centre for Gilles de la Tourette Syndrome, Groupe Hospitalier Pitié-Salpêtrière, Paris, France; Department of Psychiatry, Social psychiatry and Psychotherapy, Hannover Medical School, Hannover, Germany; Department of Psychiatry, University Medical Center Groningen & Rijksuniversiteit Groningen; MHC Drenthe Assen, the Netherlands; Institute for Anatomy and Cell Biology, Ulm University, Ulm, Germany; EMGO+ Institute for Health and Care Research, VU University Medical Centre, Amsterdam, Netherlands; Department of Child Psychiatry, Medical University of Warsaw, Warsaw, Poland; Laboratory of Neurogenetics, Department of Neurodegenerative Disorders, Mossakowski Medical Research Institute, Polish Academy of Sciences, Warsaw, Poland; Department of Molecular Biology, Institute of Biochemistry and Molecular Biology, Semmelweis University, Budapest, Hungary; Vadaskert Clinic for Child and Adolescent Psychiatry, Hungary; Department of Management, Ball State University, Muncie, IN, USA; Department of Psychiatry, Icahn School of Medicine at Mount Sinai, USA; Seaver Autism Center for Research and Treatment, Icahn School of Medicine at Mount Sinai, USA; Department of Genetics and Genomic Sciences, Icahn School of Medicine at Mount Sinai, USA; Department of Neuroscience, Icahn School of Medicine at Mount Sinai, USA; The Mindich Child Health and Development Institute, Icahn School of Medicine at Mount Sinai, USA; Friedman Brain Institute, Icahn School of Medicine at Mount Sinai, USA; Division of Tics, OCD, and Related Disorders, Icahn School of Medicine at Mount Sinai, USA; Department of Cognitive Neuroscience, Donders Institute for Brain, Cognition and Behaviour, Radboud University Medical Center, Netherlands; deCODE Genetics/Amgen, Iceland; Boehringer Ingelheim Pharma GmbH & Co. KG, CNS Research, Germany; Hematology Department- Hematopoietic Cell Transplantation Unit, Gene and Cell Therapy Center, George Papanikolaou Hospital, Greece; Department of Medicine, University of Washington, WA, USA; Altius Institute for Biomedical Sciences, WA, USA; Department of Genome Sciences, University of Washington, WA, USA; Department of Medicine, Division of Oncology, University of Washington, WA, USA; Child and Adolescent Psychiatry Department, Schneider Children’s Medical Centre of Israel, Petah-Tikva; Sackler Faculty of Medicine, Tel Aviv University, Israel; Department of Human Neurosciences, University La Sapienza of Rome, Rome, Italy; Evelina London Children’s Hospital GSTT, Kings Health Partners AHSC, London, UK; UCL Great Ormond Street Institute of Child Health, University College London, London, UK; Psychological and Mental Health Services, Great Ormond Street Hospital for Children NHS Foundation Trust, London, UK; Levvel, Academic Center for Child and Adolescent Psychiatry, Amsterdam, The Netherlands; Amsterdam UMC, Department of Child and Adolescent Psychiatry, Amsterdam, The Netherlands; Unidad de Trastornos del Movimiento. Instituto de Biomedicina de Sevilla (IBiS). Hospital Universitario Virgen del Rocío/CSIC/Universidad de Sevilla. Seville, Spain; Centro de Investigación Biomédica en Red sobre Enfermedades Neurodegenerativas (CIBERNED), Madrid, Spain; Department of Child and Adolescent Psychiatry and Psychology, Institute of Neurosciences, Hospital Clinic Universitari, Barcelona, Spain; Institut d’Investigacions Biomediques August Pi i Sunyer (IDIBAPS), Barcelona, Spain; Centro de Investigacion en Red de Salud Mental (CIBERSAM), Instituto Carlos III, Spain; Department of Psychiatry and Psychotherapy, University Hospital, LMU Munich, Munich, Germany; Institute of Systems Motor Science, University of Lübeck, Lübeck, Germany; Child and Adolescent Mental Health Centre, Mental Health Services, Capital Region of Denmark and University of Copenhagen, Copenhagen, Denmark; Division of Child and Adolescent Psychiatry, Department of Psychiatry, Lausanne University Hospital, Lausanne, Switzerland; ASL BA, Maternal and Childhood Department; Adolescence and Childhood Neuropsychiatry Unit; Bari, Italy; Department of Child and Adolescent Psychiatry, Faculty of Medicine, University Hospital Carl Gustav CarusTU Dresden, Dresden, Germany; Department of Child and Adolescent Psychiatry and Psychotherapy, University of Zurich, Zurich, Switzerland; Department of Clinical and Movement Neurosciences, UCL Queen Square, Institute of Neurology, University College London, London, UK; Department of Clinical Neurosciences, Cumming School of Medicine & Hotchkiss Brain Institute, University of Calgary, Calgary, AB, Canada; Department of Genetics and the Human Genetics Institute of New Jersey, Rutgers, the State University of New Jersey, Piscataway, NJ, USA; Department of Psychiatry and Behavioral Sciences, UCSF Weill Institute for Neurosciences, University of California, San Francisco, San Francisco, CA, USA; Quantitative Biosciences Institute, University of California, San Francisco, San Francisco, CA, USA; University of Groningen, University Medical Centre Groningen, Department of Child and Adolescent Psychiatry, Groningen, the Netherlands, Accare Child Study Center; Division of Genetic Medicine, Department of Medicine Vanderbilt University Medical Center Nashville, Nashville, TN, USA; Vanderbilt Genetics Institute, Vanderbilt University Medical Center, Nashville, TN, USA; Department of Genetics, University of North Carolina at Chapel Hill, Chapel Hill, NC, USA; Department of Clinical Neuroscience, Karolinska Institutet, Stockholm, Sweden; Department of Psychiatry, University of North Carolina at Chapel Hill, Chapel Hill, NC, USA; Department of Psychiatry and Genetics Institute, University of Florida College of Medicine, USA; Department of Neurology, Brigham and Women’s Hospital, and the Department of Neurology, Massachusetts General Hospital, Boston, MA, USA; 1st Laboratory of Medical Biology-Genetics, School of Medicine, Aristotle University of Thessaloniki, Thessaloniki, Greece

## Abstract

Tourette Syndrome (TS) is a childhood-onset neurodevelopmental disorder of complex genetic architecture, characterized by multiple motor tics and at least one vocal tic persisting for more than one year. We performed a genome-wide meta-analysis integrating a novel TS cohort with previously published data, resulting in a sample size of 6,133 TS individuals and 13,565 ancestry-matched controls. We identified a genome-wide significant locus on chromosome 5q15 and one array-wide significant locus on chromosome 2q24.2. Integration of eQTL, Hi-C and GWAS data implicated the *NR2F1* gene and associated lncRNAs within the 5q15 locus, and the *RBMS1* gene within the 2q24.2 locus. Polygenic risk scoring using previous GWAS results demonstrated statistically significant ability to predict TS status in the novel cohort. Heritability partitioning identified statistically significant enrichment in brain tissue histone marks, while polygenic risk scoring on brain volume data identified statistically significant associations with right and left putamen volumes. Our work presents novel insights in the neurobiology of TS opening up new directions for future studies.

## Introduction

Tourette Syndrome (TS) is a childhood-onset neurodevelopmental disorder characterized by multiple motor tics and at least one vocal tic persisting for more than one year [1]. The prevalence of TS is estimated in the range of 0.6-1% in school-aged children [2, 3]. It is a highly heritable disorder [4] with a population-based heritability estimated at 0.7 [5, 6] and SNP-based heritability estimates ranging from 0.21 [7] to 0.58 [4] of the total heritability. TS exhibits high polygenicity and its genetic background is influenced by both common and rare variants of small effect spread throughout the genome [4, 8, 9]. Two previously conducted genome-wide association studies (GWAS) [7, 10] have indicated enrichment of TS genetic susceptibility variants in tissues within the cortico-striatal and cortico-cerebellar circuits, and in particular, the dorsolateral prefrontal cortex [7, 10]. Furthermore, gene set analyses of GWAS data implicated ligand-gated ion channel signaling, lymphocytic, and cell adhesion and trans-synaptic signaling processes as potential biological underpinnings in the pathogenesis of TS [11]. Polygenic risk scores derived from the second TS GWAS can predict tic presence and severity at a statistically significant level [7, 12].

Here, present a genome-wide meta-analysis for TS integrating novel and previously published data resulting in a combined sample size of 6,133 TS individuals and 13,565 ancestry-matched controls. We identify a novel genome-wide significant locus in the novel (TS-EUROTRAIN) GWAS, and a second novel array-wide significant locus in the TS GWAS meta-analysis. These results provide further insight into the genetic basis of TS.

## Methods and Materials

### Datasets

The TS-EUROTRAIN dataset brings together three major TS cohorts, including 632 participants from the European Multicenter Tics in Children Study (EMTICS) [13], 763 participants from the TS-EUROTRAIN study [14], 238 participants from the TSGeneSEE study [15], and 52 participants from Sweden. These studies included participants from multiple European sites who were diagnosed using DSM-5 criteria for TS, consistent with previously published TS studies. In total, we collected samples from 1,685 individuals with TS (Supplementary Table 1). Additionally, 4,454 population control individuals were recruited from Ashkenazi Jewish, Greek, Hungarian, Polish, and Spanish sites. Ancestry-matched controls were also used from the following public datasets following appropriate approvals: British WTCCC2 1958 Birth Cohort samples (Study accession code: EGAS00000000028), German control samples obtained from the POPGEN biobank [16], and French controls from the Three City Study [17], leading to a total of 8,558 general population controls (Supplementary Table 1). Written informed consent was obtained from all participants, as approved by the ethics committees of all participating institutions.

### Genotyping, merging and imputation

Samples from the TS-EUROTRAIN dataset were genotyped on the Illumina HumanOmniExpress BeadChip. The control samples obtained from collaborators and public repositories were genotyped on multiple Illumina arrays; they were selected on the basis of maximizing marker overlap and compatibility (Supplementary Table 1). We applied standard GWAS quality control procedures to our data before and after the imputation, as described in previous GWAS performed by the Psychiatric Genomics Consortium (PGC). Quality control procedures included the removal of samples that fit any of the following criteria: call rate *<* 0.98, absolute value of inbreeding coefficient *<* 0.2, genomic sex discrepancy with reported sex, and formation of pairs with relatedness *>* 0.1875. We applied variant quality control, excluding markers with call rate *<* 0.98, differential missingness between cases and controls *<* 0.02, Hardy–Weinberg equilibrium P value *<* 1*e −* 6 in controls and *<* 1*e −* 10 in cases. Since we merged data from multiple sources, we performed the above quality control steps on each dataset separately. Imputation was performed on the Sanger Imputation Server using a reference panel of 64,940 European ancestry haplotypes (v1.1) from the Haplotype Reference Consortium (HRC) [18, 19]. We performed batch effect tests in samples of same status (case/control) between different sources, excluding markers that achieved a p-value *<* 1*e −* 5. X chromosome data were absent from a significant fraction of the external datasets obtained; it was excluded from the final analysis. As a last step, in order to avoid ancestry bias, we matched the ancestry of the controls to TS individuals, at a three to one ratio, using the first five principal components as basis. Imputation and quality control resulted in a dataset of 1,438 individuals with TS and 4,356 controls on 2,949,675 markers.

### Genome Wide Association Study

We used TS diagnosis as a categorical variable, and conducted a GWAS using an additive logistic regression model on the best guess genotypes produced by imputation. For the logistic regression we incorporated the principal components identified by Tracy-Widom statistics, as calculated by EIGENSOFT [20, 21], as well as sex and imputation batch. To control genomic inflation at SNPs with low minor allele frequency (MAF), we excluded SNPs when their MAF was <1% and minor allele count was <10, in either cases or controls. We set the level of genome-wide significance at *P* = 5*e −* 8. We used a basis of genomic distance *>* 500*kb* and linkage disequilibrium (LD) to define independence between the associated loci.

We estimated confounding bias in our results by performing LD Score Regression with *ldsc* [22] and using the attenuation ratio, as well as the p-value of the intercept to evaluate our results. Results were plotted using *matplotlib* in Python. We produced the regional plots for the top loci using the Python package *region-plot* [23], which allows the use of local LD reference panels since our data were not imputed on the 1000 Genomes reference.

### Meta-analysis

We conducted a meta-analysis with the results of the second TS GWAS study conducted by the TS Working Group of the Psychiatric Genomics Consortium (TSGWAS2) [7]. There was a known sample overlap between the TS-EUROTRAIN GWAS and TSGWAS2, and verified through genotypic identity-by-descent analysis, so the TS-EUROTRAIN GWAS was re-analyzed after excluding the overlapping samples, leading to a sample size of 1,314 cases and 4,077 ancestry-matched controls. The summary statistics of the re-analyzed TS-EUROTRAIN study and the TSGWAS2 meta-analysis were used as input to METASOFT [24]. METASOFT implements an array of methods for meta-analysis, especially in the case of heterogeneity in the results. In our study, we employed Han and Eskin’s random effects model (RE2), which separates hypothesis testing from effect-size estimation, and is demonstrated to increase statistical power under heterogeneity compared to the conventional random effects model [25, 26]. We also employed METASOFT to produce m-values, that is, estimates of the posterior probability that an effect exists, with small values indicating absence of effect, large values indicating presence of effect, and intermediate values indicating ambiguity [24]. Results were plotted using *matplotlib* and *region-plot* [23] in Python.

### Heritability and heritability partitioning

Heritability was estimated through LD Score Regression [22], after merging the alleles with the HapMap3 reference panel. We further investigated heritability partitioning into functional categories using stratified LD Score Regression [27]. TS heritability was partitioned into 53 functional categories as well as into 220 cell-type-specific and 10 cell-type-group-specific annotations produced on the data derived from the Roadmap Epigenomics Project [28]. The significance threshold for the heritability enrichments was defined at a Benjamini-Hochberg FDR *<* 0.05. P-values were calculated on the regression coefficient *τ*_*c*_.

### Genetic correlations

Bivariate LD score regression [22] was conducted to identify genetic correlations between the TS-EUROTRAIN GWAS, the TS EUROTRAIN/GWAS2 meta-analysis, and TSGWAS2 [7]. We then examined each of these studies’ cross-disorder correlations with obsessive-compulsive disorder (OCD) [29], attention deficit hyperactivity disorder (ADHD) [30], major depressive disorder (MDD) [31], autism spectrum disorders (ASD) [32], and anxiety [33]. To avoid confounding due to sample size, we selected summary statistics from studies with more than 5,000 samples. For the correlation analysis we used the European LD scores and merged alleles based on the HapMap3 reference panel for each trait, excluding markers residing in the Major Histocompatibility Complex region on chromosome 6. Significance threshold was defined by Benjamini-Hochberg false discovery rate as *<* 0.05. We visualized the results using the R packages *network-plot* and *ldsc-corr-plot* [34].

### Polygenic Risk Scoring

We used PRSice-2 [35] for our Polygenic Risk Score (PRS) analysis. We performed a unilateral PRS analysis between the TS-EUROTRAIN cohort and the TSGWAS2 [7] cohort, using the TSGWAS2 summary statistics as discovery and the TS-EUROTRAIN GWAS, after excluding the overlapping samples, as the target dataset. The TSGWAS2 summary statistics were clumped on the LD information of the TS-EUROTRAIN GWAS, using a window of 250kb and an *r*^2^ threshold of 0.1. PRSice performed the scoring on subsets of the dataset based on nine thresholds of p-value leniency (5E-08, 1E-04, 1E-03, 0.01, 0.05, 0.1, 0.2, 0.5, 1). The resulting PRS was tested for association with TS, using logistic regression with the previously mentioned ancesty components, sex, and imputation batch as covariates. The model fit for best p-value threshold was run using 10,000 permutations. Liability scale was calculated on the variance explained by the PRS (*R*^2^), using a TS population prevalence of 1%.

### Biological annotation of results

We applied FUMA [36] to perform gene-based and gene-set analyses on the results from the TS-EUROTRAIN GWAS and the subsequent TS-EUROTRAIN/GWAS2 meta-analysis. The genetic variants were assigned to protein-coding genes based on their GRCh37 build genomic position, using a *±*20kb window size. After quality control, 18,089 genes contained at least one variant and as such were used for the gene-based analysis, thus setting the Bonferroni threshold at *p <* 2.764*e −* 6. The gene-based association results were subsequently used for gene-set enrichment analysis under a competitive model. Tissue Expression Analysis was conducted on the GTEx v8 expression data [37, 38]. We investigated chromatin contact points through Capture Hi-C data available from the 3D Genome Browser [39], using promoter-centered long-range chromatin interaction data derived from human dorsolateral prefrontal cortex tissues [40].

We performed a set-based association analysis using PLINK [41, 42] on the gene-sets that were previously identified as significantly associated with TS [11, 43]. We used logistic regression as the association model on the genotypes and principal components that were identified by Tracy-Widom statistics in the GWAS. Another repetition of this step was performed with the *χ*^2^ association test, to test for this method’s robustness to population structure. We proceeded to run the analysis on all TS-EUROTRAIN samples, using a 10kb genomic window size and a million permutations. Since the permutations were performed on the phenotypic status of the samples, and only served as a method of association of the trait with the gene sets, we also corrected the results by defining the significance threshold through Bonferroni correction.

### PRS for brain anatomy

Using the TS-EUROTRAIN/GWAS2 meta-analysis summary statistics as base, we computed TS polygenic risk scores (*P RS*_*T S*_) for individuals in the UK Biobank [44] using PRSice-2 [35] and subsequently evaluated the association between risk scores and subjects’ 14 subcortical volumes (FIRST). After quality controls, 29,829 samples with brain MRI phenotypes available were included for the analysis. We assessed the association with *P RS*_*T S*_ computed using independent SNPs with meta-analyzed p-value under various thresholds (0.05, 0.001, 1e-5). For each threshold, clumping of base summary statistics was carried out under *r*^2^ = 0.1 with a *±*500kb window. Regressions between *P RS*_*T S*_ and brain volume measurements were evaluated controlling for age, sex, and the top five PCs as covariates. Bonferroni significance threshold for FIRST measurements was *p <* 1.19*e −* 3 (0.05/(14×3)).

## Results

### Mega-analysis of TS-EUROTRAIN GWAS

GWAS analysis was performed as a mega-analysis, on the combined genetic data of all collected samples, using a logistic regression model on the best-guess genotypes (genotype probability *>* 0.9) with INFO score *>* 0.9 and MAF*>* 0.01. As covariates, we included the ancestry components 1,2,4, and 5 to account for population stratification as identified by ANOVA statistics (Supplementary Table 2), sex as a binary index to account for sex stratification, and imputation batch to account for bias due to array and imputation batch effects.

The TS-EUROTRAIN GWAS identified three novel highly-correlated (*r*^2^ *>* 0.8) genome-wide significant SNPs, located near the *NR2F1 Antisense RNA 1* long non-coding RNA (*NR2F1-AS1* lncRNA) locus (Supplementary Figure 1, Figure 1a). The strongest signal was found for rs2453763 (chr5:92376460:T/A, *OR* = 0.7512, *P* = 2.62*e −* 8, MAF=0.3581), a variant 350kb upstream of *NR2F1-AS1*, and associated with decreased risk for TS. The imputation statistics for this SNP indicate high imputation quality (MAF= 0.3581, INFO= 0.99). The proximal SNPs were rs2009416 (chr5:92415111:T/C, *OR* = 0.7532, *P* = 3.31*e −* 8, MAF=0.3562) and rs1496337 (chr5:92411293:T/C, *OR* = 0.7534, *P* = 3.33*e −* 8, MAF=0.3563). Conditional analysis using the lead SNP as covariate showed no secondary signals in the region. The top (*P <* 1*e −* 5) loci detected in the novel GWAS are reported in Table 1. LD Score Regression analysis of the summary statistics did not provide evidence for genomic inflation (*λ*_*G C*_ = 1.07, intercept=1.0061, intercept p-value=0.28, attenuation ratio=0.0887).

**Table 1:** Top regions (p<1E-5) uncovered in the TS-EUROTRAIN GWAS (1,438 cases and 4,356 controls on 2,949,675 variants). One variant was identified as genome-wide significant (p<5e-8). Chromosome and region (based on hg19) are shown for index SNPs (*LD − r*^2^ *>* 0.1), as well as number of LD-associated markers in proximity (N). A1 refers to associated allele. The odds ratio (OR) and standard error (SE) are shown for the association between A1 and TS. MAF indicates the allelic frequency of allele 1 in the dataset. The reported nearest genes were determined by genomic location (*±*500 kb). The analysis was restricted to variants with MAF *≥* 0.01 and information quality (INFO) score *≥* 0.9. Chromosome X was not analyzed, since it was absent from a significant portion of the acquired datasets.

| SNP ID | Chr | P-value | A1 | OR | SE | N | Location | KB | Genes |
| --- | --- | --- | --- | --- | --- | --- | --- | --- | --- |
| rs2453763 | 5 | $2.623e-08$ | T | 0.7512 | | 25 | chr5:92322427..92559372 | 236.946 | NR2F1-AS1 |
| rs2197383 | 3 | $4.681e-07$ | A | 0.5948 | | 165 | chr3:79889497..80380401 | 490.905 | ROBO1 |
| rs3773057 | 3 | $1.388e-06$ | T | 0.1981 | | 23 | chr3:29563164..29627731 | 64.568 | RBMS3, RBMS3-AS3 |
| rs9382365 | 6 | $1.829e-06$ | G | 0.6769 | | 77 | chr6:54418351..54531232 | 112.882 | FAM83B, TINAG |
| rs152061 | 5 | $2.085e-06$ | T | 1.262 | | 196 | chr5:64778944..64989139 | 210.196 | ADAMTS6, CENPK, ERBB2IP, NLN, PPWD1, SGTB, TRAPPC13, TRIM23 |
| rs2278796 | 1 | $6.882e-06$ | T | 1.261 | | 9 | chr1:204951209..204971553 | 20.345 | CNTN2, DSTYK, NFASC, RBBP5, TMCC2, TMEM81 |
| rs34940828 | 3 | $7.531e-06$ | C | 2.104 | | 64 | chr3:123213895..123398466 | 184.572 | ADCY5, CCDC14, MYLK, MYLK-AS1, PTPLB, SEC22A |
| rs2076218 | 1 | $9.746e-06$ | A | 1.25 | | 3 | chr1:209745395..209768699 | 23.305 | C1orf74, CAMK1G, DIEXF, G0S2, HSD11B1, IRF6, LAMB3, MIR205, MIR205HG, MIR4260, TRAF3IP3 |

### Meta-analysis with TSGWAS2

The TS-EUROTRAIN GWAS was then meta-analyzed with summary statistics results from the previous largest meta-analysis of TS to date (TSGWAS2) [7] using Han and Eskin’s random effects model [24, 25]. Since there was a known sample overlap between the two studies, the TS-EUROTRAIN GWAS was re-analyzed after excluding the overlapping samples (1,314 cases and 4,077 ancestry-matched controls), with the results being very similar to the full dataset TS-EUROTRAIN GWAS (Supplementary Figure 2). Only variants overlapping in both studies were included, leading to a total of 6,133 cases, 13,565 controls and 1,955,677 variants in the final meta-analysis.

The TS-EUROTRAIN/GWAS2 meta-analysis (Figure 2) identified the three genome-wide significant SNPs of the TS-EUROTRAIN GWAS, with rs2453763 again being the top hit (chr5:92376460:T/A, random effects *P* = 4.05*e −*08) along with an array-wide significant SNP, rs10209244 (chr2:161561898:A/G, random effects *P* = 6.16*e −* 08, *M AF* = 0.01) that resides 200kb downstream of *RBMS1* (Figure 1b). The top loci detected from the meta-analysis are reported in Table 2. LD Score Regression analysis of the summary statistics did not provide evidence for genomic inflation (*λ*_*G C*_ = 1.16, intercept=1.016, intercept p-value=0.11, attenuation ratio=0.0869).

**Table 2:**
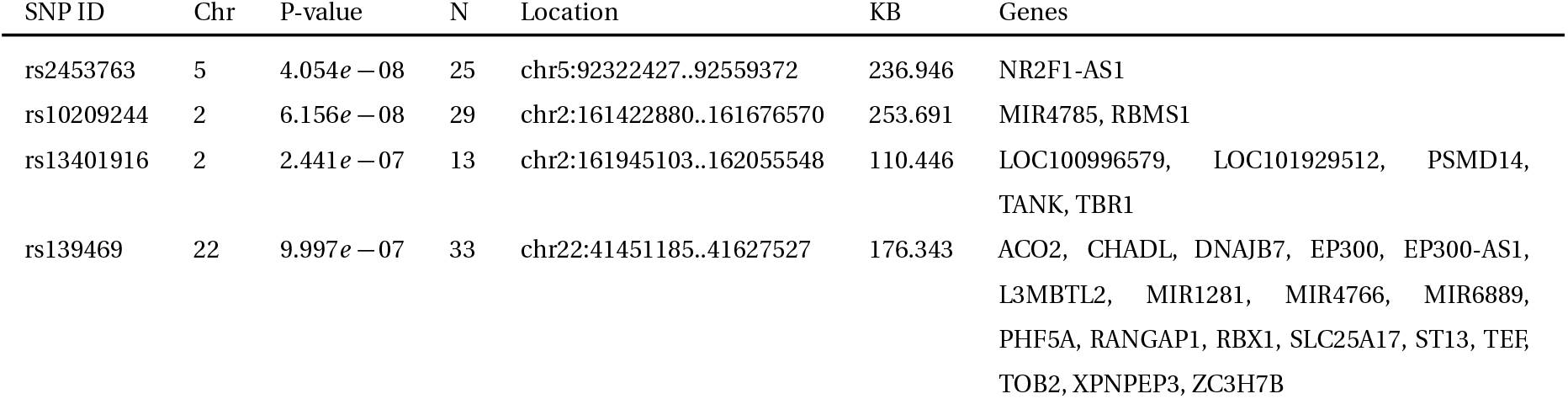
Top regions (p<1E-5) uncovered by GWAS meta-analysis (TS-EUROTRAIN and TSGWAS2 [7] - 6,133 cases, 13,565 controls on 1,955,677 variants) using Han and Eskin’s random effects model [25]. One variant was identified as genome-wide significant (p<5e-8) and one as array-wide significant (p<1e-7). Chromosome and region (based on hg19) are shown for index SNPs (LD-r2>0.1), as well as number of LD-associated markers in proximity (N). The reported nearest genes were determined by genomic location (*±*500 kb). MAF indicates the allelic frequency of the minor allele in the dataset.

**Figure 1:**
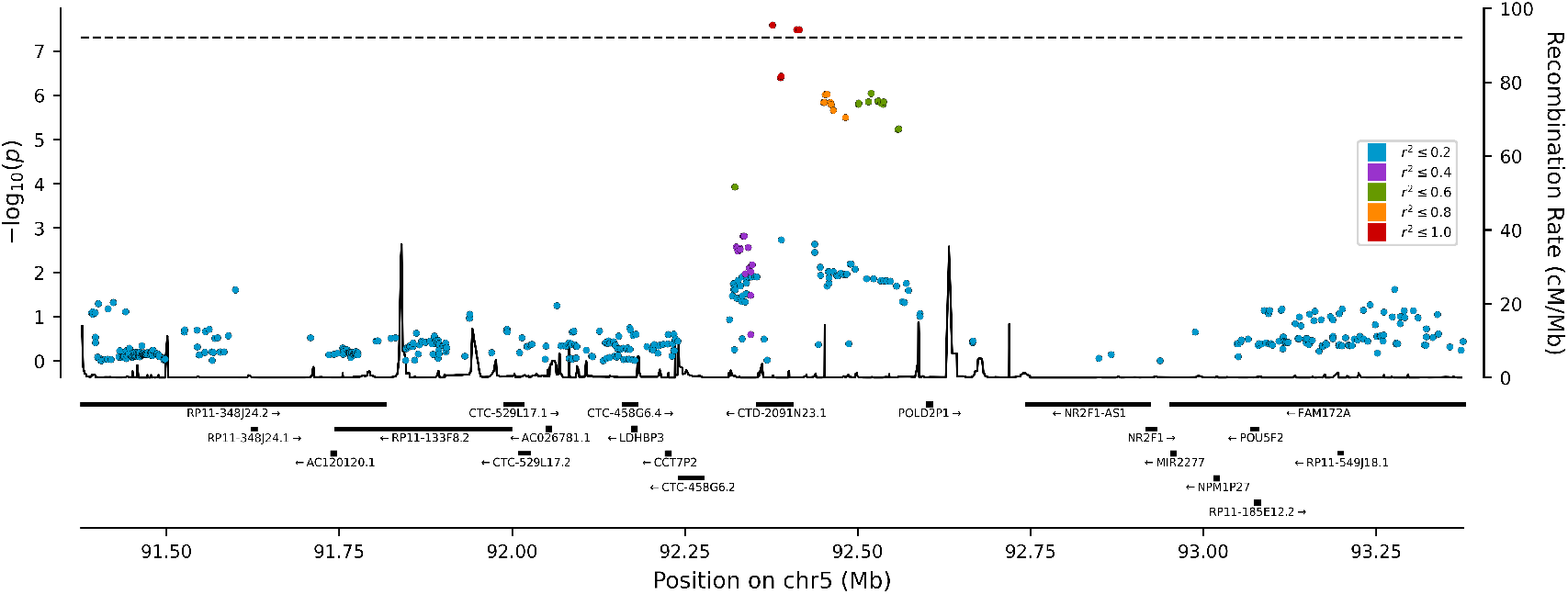
Regional plots using the TS-EUROTRAIN dataset as the base for LD calculations. In red are the markers that are in high LD with the lead marker (0.8 *≤ r*^2^ *≤* 1). a) Regional plot of the top TS-EUROTRAIN GWAS locus (NR2F1). b) Regional plot of the second top meta-analysis locus (RBMS1)

**Figure 2:**
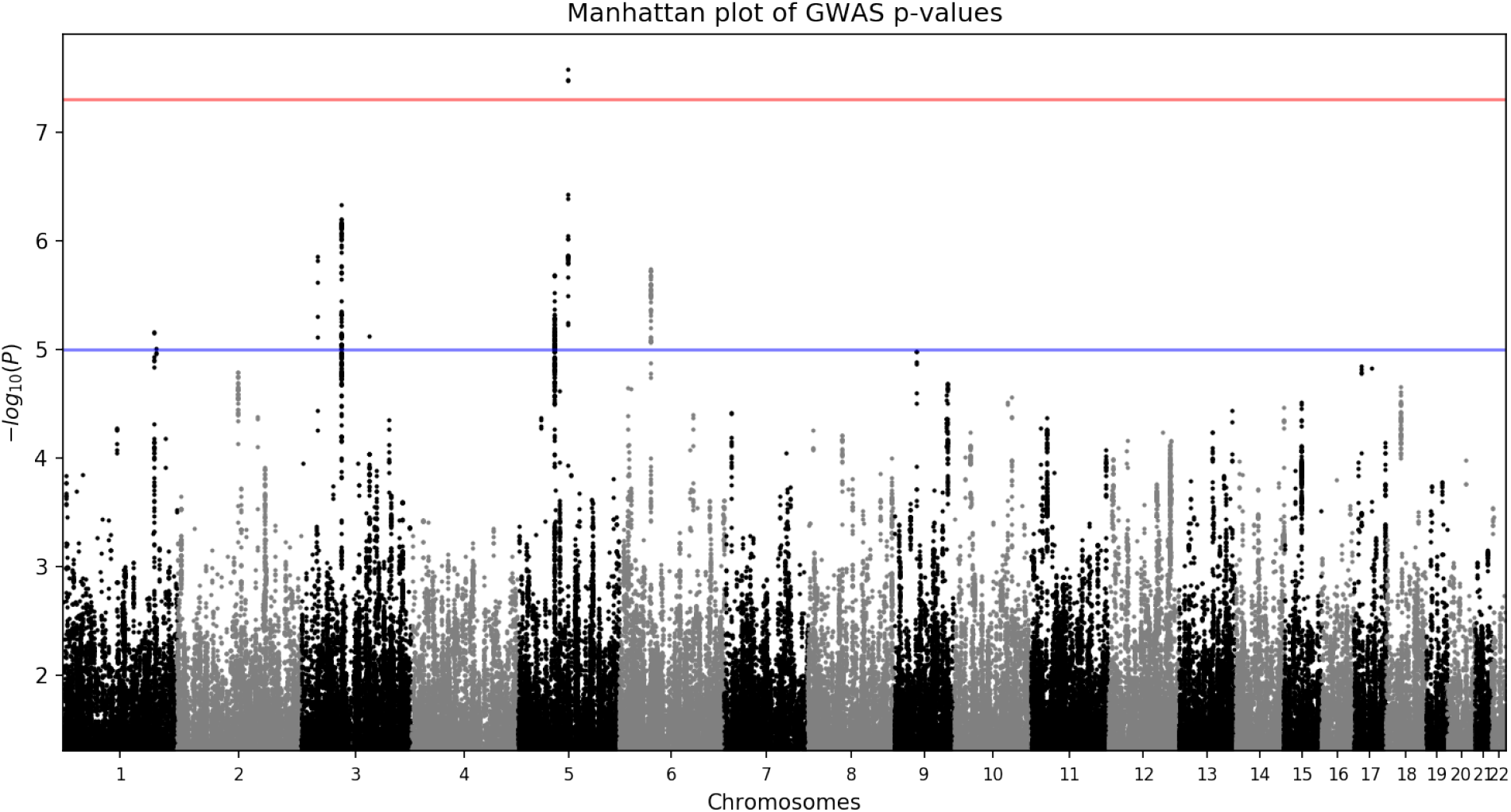
The Manhattan plot for the genome-wide association meta-analysis of Tourette Syndrome with the TS-EUROTRAIN and the TSGWAS2 datasets (6,133 TS cases and 1,3565 controls of European descent on 1,955,677 variants) using Han and Eskin’s random effects model [25]. The -log10(p) values for the association tests (two-tailed) are shown on the y axis and the chromosomes are ordered on the x axis. One genetic locus on chromosome 5 surpassed the genome-wide significance threshold (p<5e-8 ; indicated by the red line). One genetic locus surpassed the array-wide significance threshold (p<1e-7). Gray and black differentiate adjacent chromosomes.

### Genetic relationship between the TS-EUROTRAIN GWAS and TSGWAS2

The SNP with the strongest signal in the previously published TSGWAS2 study was absent from the TS-EUROTRAIN dataset due to the differences in reference panels used (1000 Genomes for TSGWAS2 and HRC for the novel study) and stringent batch effect quality control performed on the novel dataset. We observed no genome-wide significant heterogeneity (Cochran’s Q-test p-value*<* 5*e −* 8) in the meta-analysis.

To explore the relationship between the TS-EUROTRAIN GWAS and TSGWAS2, we used LD Score Regression [22] to compute their genetic correlation, after excluding the overlapping samples. LD Score Regression identified a strong genetic correlation between the two studies (*r*_*g*_ = 0.95, *p* = 6*e −* 8), and provided evidence of consistency across them (Figure 3). Investigation of the gene sets found previously associated with TS [11, 43] also successfully replicated the associations for the lymphocytic, the ligand-gated ion channel signaling, the cell adhesion and trans-synaptic signaling, as well as the astrocyte-neuron metabolic coupling gene sets (Supplementary Table 3).

**Figure 3:**
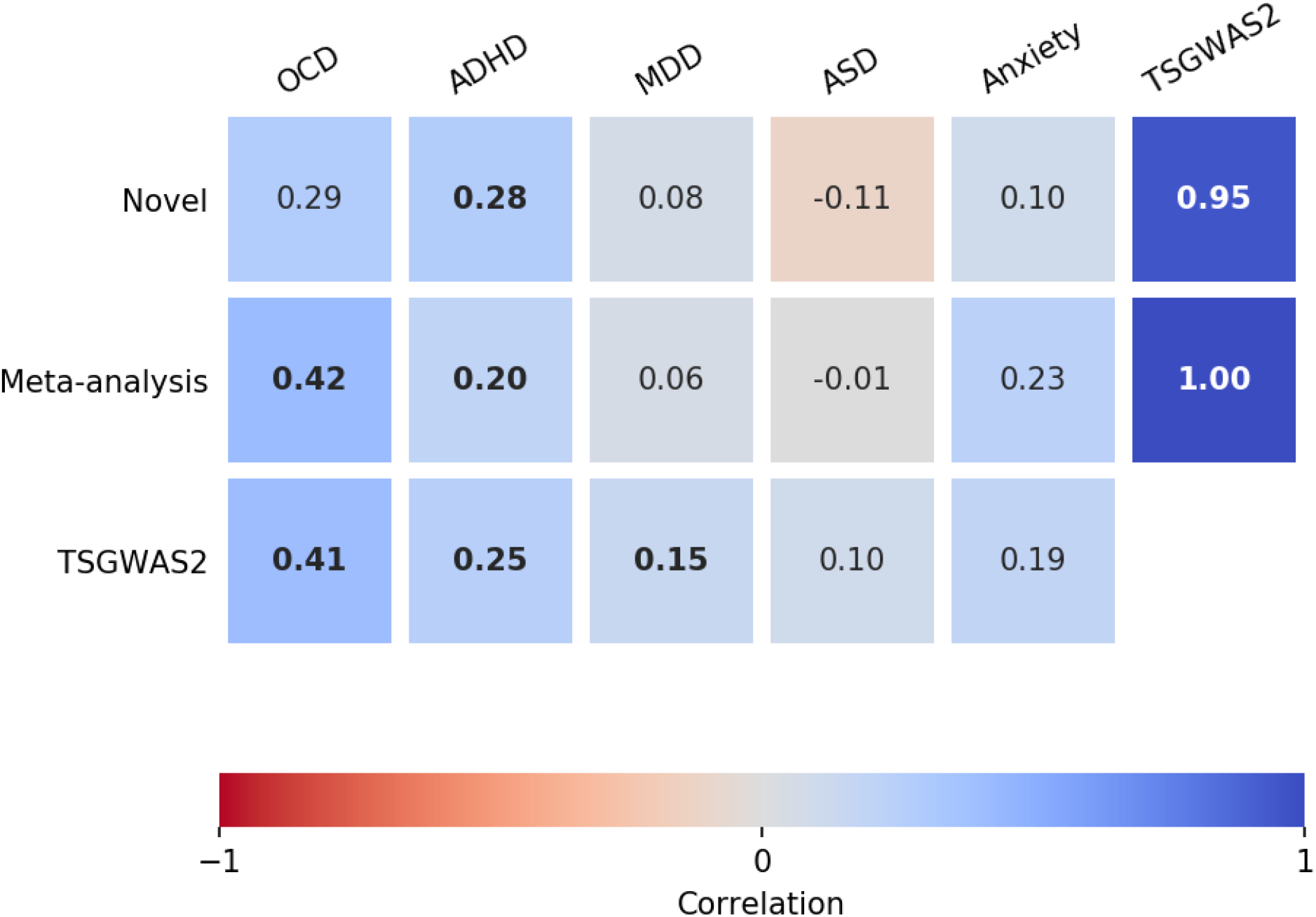
Genetic correlations with Tourette Syndrome. The genetic correlations were estimated with bivariate LD score regression [22]. We showcase the correlations between three TS studies (TS-EUROTRAIN, TS-EUROTRAIN/TSGWAS2 meta-analysis, and TSGWAS2) and five neuropsychiatric disorders (OCD, ADHD, MDD, ASD, and anxiety) previously correlated with TS. The number in each square denotes the correlation rg. In bold are the correlations that were identified as statistically significant after Benjamini-Hochberg FDR correction (a=0.05).

PRS analysis displayed consistency between the two studies. PRS were computed using the summary statistics of TSGWAS2 as a training dataset and the TS-EUROTRAIN raw genotypes as discovery in PRSice [45]. The best fit p-value threshold was determined at *p* = 0.1182 (model fit *p* = 1.26*e −* 28) (Figure 4). Maximum variance explained at the best fit model was estimated by Nagelkerke’s *R*^2^ at 3.3%.

**Figure 4:**
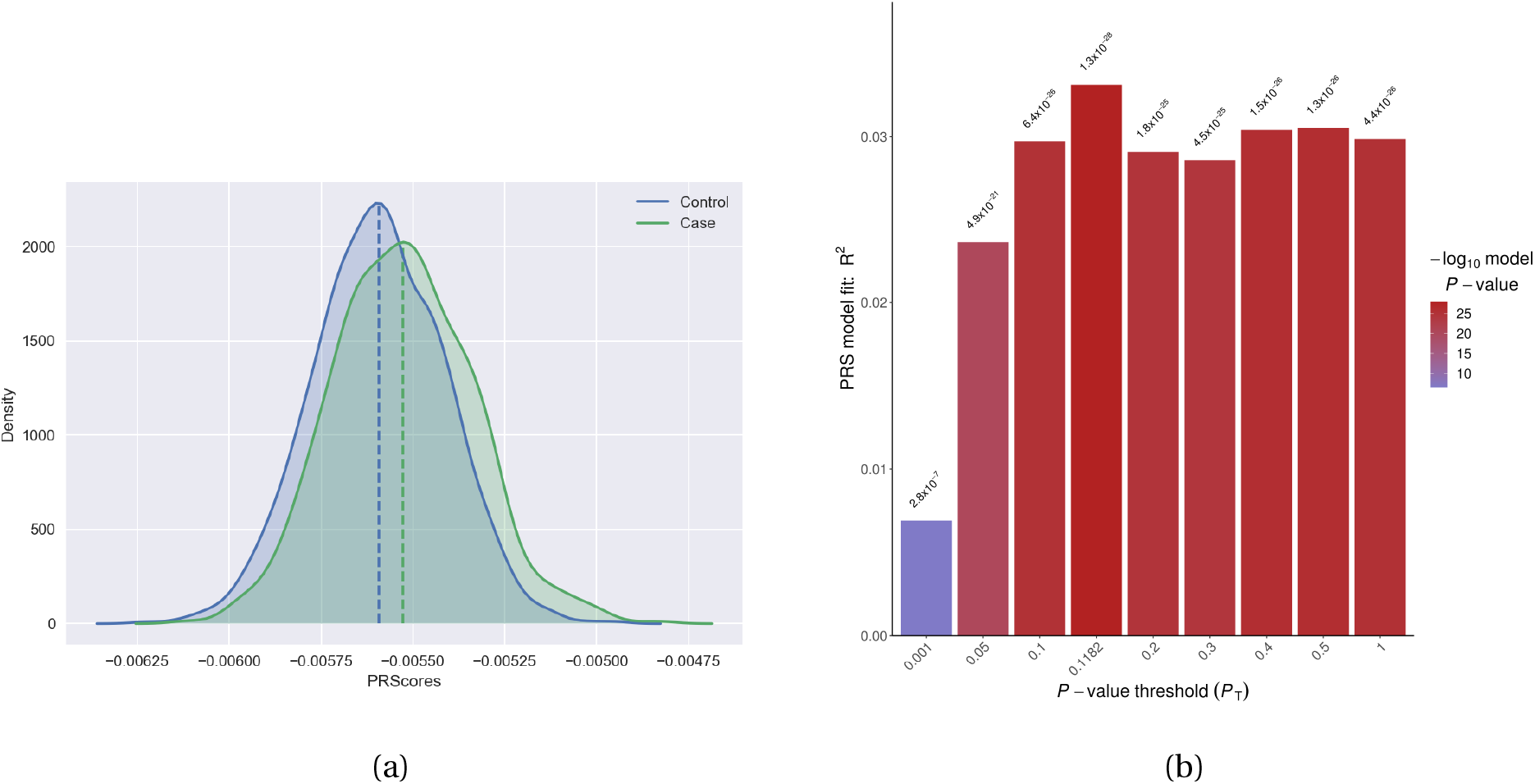
Polygenic Risk Scoring analysis using the TSGWAS2 dataset [7] as discovery and the TS-EUROTRAIN dataset as target. Best fit p-value threshold was determined at p=0.1182 (model fit p=1.26e-28). Maximum variance explained at the best fit model was estimated by Nagelkerke’s R2 at 3.3%. a) PRS distribution comparison between cases and controls for the best fit model. b) PRS histogram for each p-value bin, including the best fit bin.

### Cross-disorder analysis

Pairwise genetic correlations were computed between our results and five traits that have been found to be correlated in previous studies with the TSGWAS2 results [46, 47] using LD Score Regression [22]. Benjamini-Hochberg FDR correction with an *α* = 0.05 was used to correct for multiple testing. After correction, the TS-EUROTRAIN GWAS was significantly correlated with ADHD (*r*_*g*_ = 0.28, *p* = 7.7*e −* 3), and the TS-EUROTRAIN/GWAS2 meta-analysis was significantly correlated with ADHD (*r*_*g*_ = 0.19, *p* = 5.1*e −* 3) and OCD (*r*_*g*_ = 0.42, *p* = 1*e −* 4) (Figure 3).

### Heritability estimation and partitioning

We used LD Score Regression [22] to estimate the SNP-based heritability 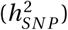 using the summary statistics of the novel GWAS and the meta-analysis. The summary statistics were merged with the HapMap3 marker panel provided by the authors. For the TS-EUROTRAIN GWAS, analysis yielded an observed scale 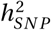 estimate of 0.4385(*SE* : 0.1167) on the observed scale, and 0.2736(*SE* : 0.0728) assuming a TS prevalence of 0.01 on the liability scale, while the LD score regression analysis intercept was computed at 1.0157(*SE* : 0.013) (p-value 0.028) and the ratio of stratification to polygenicity was estimated at 0.0863(*SE* : 0.0711). For the meta-analysis the 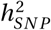 estimate was 0.3504(*SE* : 0.0439) and 0.2184(*SE* : 0.0269) on the liability scale.

We proceeded to partition the heritability by functional genomic categories using stratified LD Score Regression [27] on the full baseline model and a model based on the Roadmap epigenomics data, as provided by the authors [27]. The full baseline model included 24 main overlapping functional categories and identified statistically significant enrichment in two categories, after Benjamini-Hochberg FDR correction at an *α* = 0.05. The H3K4me1 sites category (enrichment value 1.61, *P* = 9.5*e −*4) was the top significant signal in the analysis, with the conserved elements category (enrichment value 2.05,*P* = 3.8*e −* 3) being the second significant signal. The Roadmap model includes epigenomic mapping data from 395 tissues [28] and when applied to our data for heritability partitioning, yielded 13 statistically significant modifications after Benjamini-Hochberg FDR correction at an *α* = 0.05. These 13 signals marked the enrichment of the histone marks H3K27ac, H3K4me1, and H3K9ac on five brain tissues. H3K27ac was identified on the angular gyrus, the cingulate gyrus, the dorsolateral prefrontal cortex, and the inferior temporal lobe; H3K4me1 on the angular gyrus, the cingulate gyrus, the dorsolateral prefrontal cortex, the inferior temporal lobe, and the substantia nigra; H3K9ac on the angular gyrus, the anterior caudate, the dorsolateral prefrontal cortex, and the inferior temporal lobe. The results for the full baseline model and the Roadmap model can be further explored in Supplementary Tables 4 and 5.

### Biological annotation and enrichment analysis

Functional mapping, annotation, and gene set enrichment using the FUMA pipeline did not produce significant results. The identified top signals from the TS-EUROTRAIN GWAS and the TS-EUROTRAIN/GWAS2 meta-analysis reside in large intergenic regions whose distance from their correlated genes (Supplementary Figure 5) exceeded the distance limits set by the software, and were thus excluded from the annotation step of the pipeline. The top signal of the gene-based analysis was *RANGAP1* (*P* = 3.36*e −* 6) on chromosome 22; it did not meet the genome-wide significance threshold (*P* = 2.8*e −* 6) (Supplementary Table 6). MAGMA tissue expression analysis using FUMA did not produce any statistically significant results. MAGMA tissue expression analysis on the TS-EUROTRAIN GWAS using the 53 GTEx tissue sample set indicated putative enrichment in various brain tissues, with the top signals in the hypothalamus, putamen, and nucleus accumbens. Analysis using the 30 GTEx tissue sample set indicated potential enrichment in the brain, followed by the colon, adrenal gland, and pituitary (Supplementary Figure 3). MAGMA tissue expression analysis of the meta-analysis, using the 53 tissue sample set from GTEx, indicated stronger putative enrichment in various brain tissues, with the top signals in the cerebellar hemisphere, cerebellum, and frontal cortex (area BA9). Using the 30 tissue sample set from GTEx, stronger evidence of potential enrichment could be identified in the brain, followed by the pituitary and the ovary (Supplementary Figure 4).

### Brain volumes associated through PRS

Association between the meta-analysis *PRS*_*T S*_ and brain volume measurements highlighted the previously described relationship [48] between genetic risk for TS and bilateral putamen volumes under multiple PRS p-value thresholds. The strongest (and only statistically significant) associations were found between *PRS*_*T S*_ computed using 116 independent SNPs with *p <* 0.001 and right putamen volume (PRS-p = 3.70E-05, *r*^2^ = 0.264), as well as left putamen volume (PRS-p = 1.25E-04, *r*^2^ = 0.257), both being negatively associated with TS genetic risk (Supplementary Table 7).

## Discussion

We report results from a novel study, as well as the largest GWAS meta-analysis on TS to date, including 6,133 TS individuals of European ancestry and 13,844 matched controls. We report two novel independent genome-wide significant loci associated with TS: one on chromosome 5q15 upstream of *NR2F1* in TS-EUROTRAIN GWAS (1,438 TS cases and 4,356 controls) and another on chromosome 2q24.2 downstream of *RBMS1* in the final meta-analysis. These results confirm the value of collaborative efforts towards expanding sample size and datasets available for analysis (such as the TS-EUROTRAIN, EMTICS, TSGeneSEE, and PGC initiatives). However, this study still only captures a small fraction of the risk for TS attributable to common variants. Larger studies are necessary and warranted.

The genome-wide significant locus identified in the TS-EUROTRAIN/GWAS2 meta-analysis resides in the 5q15 chromosomal region (SNP rs2453763). The associated SNP is located within *CTD2091N23*.*1*, a gene encoding a long non-coding RNA that has yet to be functionally characterized. It is found upstream of a gene cluster that harbors the genes *NR2F1-AS, NR2F1*, and *lnc-NR2F1*. The GTEx portal [38] reports SNP rs2453763 to be significantly associated as an splicing quantitative trait locus (sQTL) for *CTD-2091N23*.*1* in the tibial nerve, and as an eQTL for *NR2F1* and *NR2F1-AS* in the esophagus smooth muscles and for *CTD-2091N23*.*1* in cultured fibroblasts (Table 3). Capture Hi-C [39] showed strong evidence for the SNP being related to the regulation of *NR2F1* (Supplementary Figure 5).

**Table 3:**
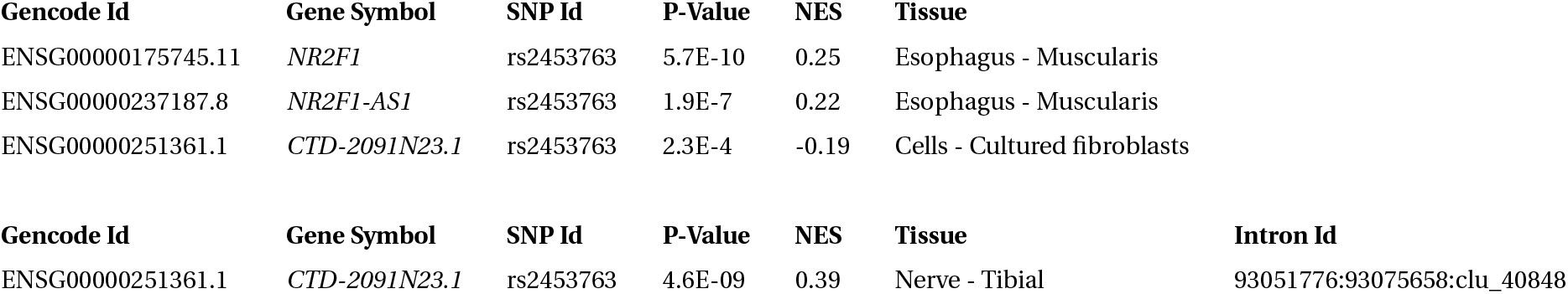
Significant SNP-gene pairings identified through GTEx eQTL and sQTL data [37, 38]. Four significant associations were identified for SNP rs2453763, while no significant associations were identified for rs10209244. rs2453763 is an eQTL for three genes on two tissues, and an sQTL for one gene on one tissue. Reported are the symbol of the associated gene, the respective associated tissue, and the normalized effect size (NES). a) GTEx eQTL associations for the top variant in *NR2F1* (rs2453763). b) GTEx sQTL associations for the top variant in *NR2F1* (rs2453763)

Moreover, the 5q15 region has previously been implicated in neurodevelopmental phenotypes [49–52]. The 5q14-5q15 regions have been reported to contain fragile sites that are associated with genomic and epigenomic instability as well as linked to schizophrenia and autism [53]. The exact genes are yet to be identified, with recent evidence suggesting a role for *NR2F1*-related genes, and more intriguingly, the *lnc-NR2F1* gene. *lnc-NR2F1* is a long non-coding RNA locus discovered to be recurrently mutated in individuals with autism spectrum disorders and intellectual disability, with translocations in this locus reported to show patterns of Mendelian inheritance [54]. A functional study of *lnc-NR2F1* identified its role in neuronal maturation *in vitro* through expression regulation of a network of genes that have been linked to autism [54]. Functional studies of the *NR2F1* gene also have indicated its critical role for neurodevelopment through investigations into human and mouse haploinsufficiency [55], insertion of point mutations in mouse models that lead to excitatory/inhibitory neuronal imbalance [56], and the study of knock-out mouse models [57]. Notably, in the absence of *NR2F1*, an imbalance between oligodendrocytes and astrocytes develops, leading to postnatal hypomyelination and astrogliosis [55]. NR2F1 is a highly conserved orphan nuclear receptor which is a regulator of transcription. It belongs to the steroid/thyroid hormone nuclear receptor superfamily, involved in a wide range of roles, including cell differentiation, cancer progression, and central and peripheral neurogenesis [58]. A multitude of pathogenic variants have been identified in NR2F1, leading to Bosch-Boonstra-Schaaf optic atrophy syndrome, and autosomal dominant neurodevelopmental disorder (OMIM: 615722) [59]. *NR2F1* is also known by its historical name, *COUPTF1*; it is a target of the androgen receptor (AR), along with *SOX9* and *OCT4* [60]. NR2F1 interacts with SOX9 [60, 61] and represses a host of targets in multiple tissues, including *CYP17A1*, Oxytocin gene *OXT*, and *OCT4* [62]. Especially in the case of *CYP17A1*, encoding for a key enzyme of steroid biosynthesis, NR2F1 and SF-1 exert opposing effects, as repressor and activator, respectively [63].

The second locus (revealed by the meta-analysis) was located in the 2q24.2 chromosomal region (SNP rs10209244), downstream of the *RBMS1* gene. Capture Hi-C [39] showed strong evidence for the SNP being related to the regulation of *RBMS1* (Supplementary Figure 5). In previous GWAS, *RBMS1* has been associated with lymphocyte, neutrophil, and white blood cell count [64, 65], educational attainment and mathematical ability [66], and addiction to tobacco and alcohol [67]. RBMS1 has been implicated in estradiol production on granulosa cells, through inactivation of c-Myc, which is its downstream target [68, 69]. Specifically, *RBMS1* mRNA stability is controlled through miR-383, which is in turn positively regulated by SF-1 [68, 69]. These results hint that steroid regulatory pathways may be involved in TS pathogenesis. Steroid hormones have been proposed to play a fundamental role in TS and sexual dimorphism of the central nervous system; androgenic hormones, in particular, are likely to exacerbate symptoms of the disorder [70].

We sought to validate our results through means of heritability correlation patterns and polygenic risk scoring. Heritability analysis in the TS-EUROTRAIN dataset indicated that a large proportion of TS SNP-based heritability can be attributed to common variants 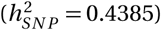, in concordance with the estimate ranges in previous investigations [4, 7]. The attenuation ratio was sufficiently low at 0.0887, attributing inflation to polygenicity in the samples. After exclusion of the overlapping samples, heritability correlation between the previous TS GWAS and the TS-EUROTRAIN dataset indicated an almost complete correlation between the two studies. Polygenic prediction in the TS-EUROTRAIN cohort using the TSGWAS2 results as discovery achieved significant predictive levels, on par with the inter-cohort predictive Nagelkerke’s *R*^2^ levels in the previous TS GWAS [7] and substantially increased by more than an order of magnitude compared to tic prediction in a general population cohort [12].

Investigations into the genetic basis of TS have long been hampered by heterogeneity between studies. We addressed this by employing a random-effects meta-analytic method, Han and Eskin’s random effects model [25], a model shown to perform particularly well in detecting true positives in the presence of heterogeneity in complex disorders [26].

The neuroendocrine system has long been hypothesized to be involved in the neurobiology of TS [71]. The hypothalamus-pituitary-gonadal (HPG) axis has been hypothesized to be implicated in TS tic exacerbation [71], while a series of investigations have been launched into deciphering the role of stress through the hypothalamus-pituitary-adrenal (HPA) axis [72, 73]. The FUMA eQTL analysis did not provide statistically significant results, however, it demonstrated distinct patterns of enrichment towards brain tissues, the pituitary, and the ovary when performed on the meta-analysis results, and towards the adrenal glands, the brain, and the pituitary when performed on the TS-EUROTRAIN GWAS results. While future analyses will be needed to replicate these findings, these results might suggest the potential involvement of the HPG and the HPA axes in TS pathogenesis.

In summary, our GWAS meta-analysis of 6,133 cases and 13,565 controls identified a genomewide significant locus on chromosome 5q15 and one array-wide significant locus on chromosome 2q24.2. Integration of eQTL and GWAS data implicate the *NR2F1* gene and associated lncRNAs within the 5q15 locus, and Hi-C data provides some evidence for the RBMS1 gene within the 2q24.2 locus. While this study is, to our knowledge, the largest TS GWAS meta-analysis to date, when compared to GWAS in other neuropsychiatric disorders, it is clear that even larger studies are warranted. Increased statistical power will further enable the identification of more leads for potential future interventions. Future plans of collaborative efforts will focus on vastly reinforcing sample sizes, increasing power to adequately perform fine-mapping and eQTL analyses with the aim to move towards the elucidation of the underlying biology of TS.

## Data Availability

Summary statistics available online.

https://github.com/ftsetsos/tseurotraingwas2021

## Funding

This study was supported by the EMTICS (FP7-HEALTH, Grant agreement ID: 278367), TS-EUROTRAIN (FP7-PEOPLE, Grant agreement ID: 316978), and NINDS (1R01NS105746-01A1) grants. AJW was funded by R01NS105746, the Tourette Association of America, and the Weill Institute for Neurosciences. AM was funded by the Deutsche Forschungsgemeinschaft (DFG; FOR 2698). AS received support from the NIHR UCL/H Biomedical Research Centre. BH is an employee of Boehringer Ingelheim Pharma. CJ was funded by Lundbeck Fonden, grant number R100-2011-9332. CB was supported by funding from the Merit-prize fellowship of Semmelweis University, the Bolyai Janos research fellowship of the Hungarian Academy of Sciences BO/00987/16/5, the UNKP-18-4 of the new National Excellence Program of the Ministry of Human Capacities and the Baron Munchausen Program of the Institute of Medical Chemistry, Molecular Biology and Pathobiochemistry, Semmelweis University. DCC was funded by the TSAA, by the Stichting VC-GGZ, and by TS-EUROTRAIN. DM has received research support from Ipsen Corporate and funding grants from Dystonia Medical Research Foundation Canada, Parkinson Canada, The Owerko Foundation, and the Michael P Smith Family. LKD was supported by grants from the National Institutes of Health including R01NS102371 and R01NS105746. PM has received grants from the Spanish Ministry of Science and Innovation [RTC2019-007150-1], the Instituto de Salud Carlos III-Fondo Europeo de Desarrollo Regional (ISCIIIFEDER), PI16/01575, PI19/01576], the Consejeria de Economia, Innovacion, Ciencia y Empleo de la Junta de Andalucia [CVI-02526, CTS-7685], the Consejeria de Salud y Bienestar Social de la Junta de Andalucia, PE-0210-2018]. PJ and CZ were funded by the National Science Center, Poland: UMO2016/23/B/NZ2/03030. ZT funded by Lundbeck Fonden, grant number R100-2011-9332. TIC Genetics was supported by grants from NIH (MH115958, MH115960, MH115962, MH115961, MH115993, MH115963, MH115959) and the New Jersey Center for Tourette Syndrome and Associated Disorders (NJCTS).

## Acknowledgements

The TSAICG includes Cathy L. Barr, James R. Batterson, Cheston Berlin, Cathy L. Budman, Giovanni Coppola, Nancy J. Cox, Sabrina Darrow, Yves Dion, Nelson B. Freimer, Marco A. Grados, Erica Greenberg, Matthew E. Hirschtritt, Alden Y. Huang, Cornelia Illmann, Robert A. King, Roger Kurlan, James F. Leckman, Gholson J. Lyon, Irene A. Malaty, William M. McMahon, Benjamin M. Neale, Michael S. Okun, Lisa Osiecki, Mary M. Robertson, Guy A. Rouleau, Paul Sandor, Harvey S. Singer, Jan H. Smit, Jae Hoon Sul

The TSGeneSEE initiative includes Christos Androutsos, Entela Basha, Luca Farkas, Jakub Fichna, Piotr Janik, Mira Kapisyzi, Iordanis Karagiannidis, Anastasia Koumoula, Peter Nagy, Joanna Puchala, Natalia Szejko, Urszula Szymanska, Vaia Tsironi

The EMTICS collaborative group includes Alan Apter, Juliane Ball, Benjamin Bodmer, Emese Bognar, Judith Buse, Marta Correa Vela, Carolin Fremer, Blanca Garcia-Delgar, Mariangela Gulisano, Annelieke Hagen, Julie Hagstrøm, Marcos Madruga-Garrido, Peter Nagy, Alessandra Pellico, Daphna Ruhrman, Jaana Schnell, Paola Rosaria Silvestri, Liselotte Skov, Tamar Steinberg, Friederike Tagwerker Gloor, Victoria L. Turner, Elif Weidinger

The TS-EUROTRAIN network includes John Alexander, Tamas Aranyi, Wim R. Buisman, Jan K. Buitelaar, Nicole Driessen, Petros Drineas, Siyan Fan, Natalie J. Forde, Sarah Gerasch, Odile A. van den Heuvel, Cathrine Jespersgaard, Ahmad S. Kanaan, Harald E. Möller, Muhammad S. Nawaz, Ester Nespoli, Luca Pagliaroli, Geert Poelmans, Petra J. W. Pouwels, Francesca Rizzo, Dick J. Veltman, Ysbrand D. van der Werf, Joanna Widomska, Nuno R. Zilhäo The TIC Genetics authors collaborative group includes Lawrence W. Brown, Keun-Ah Cheon, Barbara J. Coffey, Thomas V. Fernandez, Blanca Garcia-Delgar, Donald L Gilbert, Julie Hagstrøm, Hyun Ju Hong, Laura Ibanez-Gomez, Eun-Joo Kim, Young Key Kim, Young-Shin Kim, Robert A. King, Yun-Joo Koh, Sodahm Kook, Samuel Kuperman, Bennett L. Leventhal, Marcos Madruga-Garrido, Athanasios Maras, Tara L. Murphy, Eun-Young Shin, Dong-Ho Song, Jungeun Song, Matthew W State, Frank Visscher, Sheng Wang, Samuel H. Zinner

## Notes

### Competing Interest Statement

The authors have declared no competing interest.

### Funding Statement

This study was supported by the EMTICS (FP7-HEALTH, Grant agreement ID: 278367), TSEUROTRAIN (FP7-PEOPLE, Grant agreement ID: 316978), and NINDS (1R01NS105746-01A1) grants. AJW was funded by R01NS105746, the Tourette Association of America, and the Weill Institute for Neurosciences. AM was funded by the Deutsche Forschungsgemeinschaft (DFG; FOR 2698). AS received support from the NIHR UCL/H Biomedical Research Centre. BH is an employee of Boehringer Ingelheim Pharma. CJ was funded by Lundbeck Fonden, grant number R100-2011-9332. CB was supported by funding from the Merit-prize fellowship of Semmelweis University, the Bolyai Janos research fellowship of the Hungarian Academy of Sciences BO/00987/16/5, the UNKP-18-4 of the new National Excellence Program of the Ministry of Human Capacities and the Baron Munchausen Program of the Institute of Medical Chemistry, Molecular Biology and Pathobiochemistry, Semmelweis University. DCC was funded by the TSAA, by the Stichting VC-GGZ, and by TS-EUROTRAIN. DM has received research support from Ipsen Corporate and funding grants from Dystonia Medical Research Foundation Canada, Parkinson Canada, The Owerko Foundation, and the Michael P Smith Family. LKD was supported by grants from the National Institutes of Health including R01NS102371 and R01NS105746. PM has received grants from the Spanish Ministry of Science and Innovation [RTC2019-007150-1], the Instituto de Salud Carlos III-Fondo Europeo de Desarrollo Regional (ISCIII-FEDER), PI16/01575, PI19/01576], the Consejeria de Economia, Innovacion, Ciencia y Empleo de la Junta de Andalucia [CVI-02526, CTS-7685], the Consejeria de Salud y Bienestar Social de la Junta de Andalucia, PE-0210-2018]. PJ and CZ were funded by the National Science Center, Poland: UMO-2016/23/B/NZ2/03030. ZT funded by Lundbeck Fonden, grant number R100-2011-9332. TIC Genetics was supported by grants from NIH (MH115958, MH115960, MH115962, MH115961, MH115993, MH115963, MH115959) and the New Jersey Center for Tourette Syndrome and Associated Disorders (NJCTS).

### Author Declarations

The study was ruled as exempt category 4 (secondary research of non-identifiable data) by the Purdue University IRB

## References

1. Robertson MM, Cavanna AE, Eapen V. Gilles de la Tourette syndrome and disruptive behavior disorders: prevalence, associations, and explanation of the relationships. The Journal of neuropsychiatry and clinical neurosciences 27, 33–41. doi:10.1176/appi.neuropsych.13050112 (2015).

2. Scharf JM, Miller LL, Gauvin CA, Alabiso J, Mathews CA, Ben-Shlomo Y. Population prevalence of Tourette syndrome: A systematic review and meta-analysis. Movement Disorders 30, 221–228. doi:10.1002/mds.26089 (2015).

3. Robertson MM, Eapen V, Cavanna AE. The international prevalence, epidemiology, and clinical phenomenology of Tourette syndrome: A cross-cultural perspective. Journal of Psychosomatic Research 67, 475–483. doi:10.1016/j.jpsychores.2009.07.010 (2009).

4. Davis LK et al. Partitioning the Heritability of Tourette Syndrome and Obsessive Compulsive Disorder Reveals Differences in Genetic Architecture. PLoS Genetics. doi:10.1371/journal.pgen.1003864 (2013).

5. Robertson MM et al. Gilles de la Tourette syndrome. Nature Reviews Disease Primers 3, 16097. doi:10.1038/nrdp.2016.97 (2017).

6. Mataix-Cols D et al. Familial Risks of Tourette Syndrome and Chronic Tic Disorders: A Population-Based Cohort Study. JAMA psychiatry 72, 787–793. doi:10.1001/jamapsychiatry.2015.0627 (2015).

7. Yu D* et al. Interrogating the genetic determinants of Tourette syndrome and other tic disorders through genome-wide association studies. American Journal of Psychiatry (2018).

8. Huang A et al. Rare Copy Number Variants in NRXN1 and CNTN6 Increase Risk for Tourette Syndrome. Neuron 94. doi:10.1016/j.neuron.2017.06.010 (2017).

9. Wang S et al. De Novo Sequence and Copy Number Variants Are Strongly Associated with Tourette Disorder and Implicate Cell Polarity in Pathogenesis. Cell Reports 24, 3441–3454.e12. doi:10.1016/j.celrep.2018.08.082 (2018).

10. Scharf JM et al. Genome-wide association study of Tourette’s syndrome. Molecular psychiatry 18, 721–8. doi:10.1038/mp.2012.69 (2013).

11. Tsetsos F et al. Synaptic processes and immune-related pathways implicated in Tourette syndrome. Translational Psychiatry 11. doi:10.1038/s41398-020-01082-z (2021).

12. Abdulkadir M et al. Polygenic Risk Scores Derived From a Tourette Syndrome Genome-wide Association Study Predict Presence of Tics in the Avon Longitudinal Study of Parents and Children Cohort. Biological Psychiatry 85. Neurodevelopmental Alterations and Childhood Behavior, 298–304. doi:https://doi.org/10.1016/j.biopsych.2018.09.011 (2019).

13. Schrag A et al. European Multicentre Tics in Children Studies (EMTICS): protocol for two cohort studies to assess risk factors for tic onset and exacerbation in children and adolescents. European Child & Adolescent Psychiatry 28, 91–109. doi:10.1007/s00787-018-1190-4 (2019).

14. Forde NJ et al. TS-EUROTRAIN: A European-wide investigation and training network on the etiology and pathophysiology of Gilles de la Tourette Syndrome. Frontiers in Neuroscience 10. doi:10.3389/fnins.2016.00384 (2016).

15. Karagiannidis I et al. Replication of association between a SLITRK1 haplotype and Tourette Syndrome in a large sample of families. Molecular Psychiatry 17, 665–668. doi:10.1038/mp.2011.151 (2012).

16. Nöthlings U, Krawczak M. PopGen: Eine populationsbasierte Biobank mit Langzeitverfolgung der Kontrollkohorte. Bundesgesundheitsblatt Gesundheitsforschung - Gesundheitsschutz 55, 831–835. doi:10.1007/s00103-012-1487-2 (2012).

17. Alpérovitch A et al. Vascular factors and risk of dementia: Design of the Three-City Study and baseline characteristics of the study population. Neuroepidemiology 22, 316–325. doi:10.1159/000072920 (2003).

18. Das S et al. Next-generation genotype imputation service and methods. Nature Genetics 48, 1284–1287. doi:10.1038/ng.3656 (2016).

19. Consortium tHR et al. A reference panel of 64,976 haplotypes for genotype imputation. Nature Genetics 48, 1279–1283. doi:10.1038/ng.3643 (2016).

20. Price AL, Patterson NJ, Plenge RM, Weinblatt ME, Shadick NA, Reich D. Principal components analysis corrects for stratification in genome-wide association studies. Nature Genetics 38, 904–909. doi:10.1038/ng1847 (2006).

21. Patterson N, Price AL, Reich D. Population structure and eigenanalysis. PLoS Genetics 2, 2074–2093. doi:10.1371/journal.pgen.0020190 (2006).

22. Bulik-Sullivan BK et al. LD Score regression distinguishes confounding from polygenicity in genome-wide association studies. Nature Genetics 47, 291–295. doi:10.1038/ng.3211 (2015).

23. Tardif JC et al. Pharmacogenomic Determinants of the Cardiovascular Effects of Dalcetrapib. Circulation: Cardiovascular Genetics 8, 372–382. doi:10.1161/CIRCGENETICS.114.000663 (2015).

24. Han B, Eskin E. Interpreting meta-analyses of genome-wide association studies. PLoS Genetics 8, 1002555. doi:10.1371/journal.pgen.1002555 (2012).

25. Han B, Eskin E. Random-effects model aimed at discovering associations in meta-analysis of genome-wide association studies. American Journal of Human Genetics 88, 586–598. doi:10.1016/j.ajhg.2011.04.014 (2011).

26. Urbut SM, Wang G, Carbonetto P, Stephens M. Flexible statistical methods for estimating and testing effects in genomic studies with multiple conditions. Nature Genetics 51, 187–195. doi:10.1038/s41588-018-0268-8 (2019).

27. Finucane HK et al. Partitioning heritability by functional annotation using genome-wide association summary statistics. Nature Genetics 47, 1228–1235. doi:10.1038/ng.3404 (2015).

28. Kundaje A et al. Integrative analysis of 111 reference human epigenomes. Nature 518, 317–330. doi:10.1038/nature14248 (2015).

29. Arnold PD et al. Revealing the complex genetic architecture of obsessive–compulsive disorder using meta-analysis. Molecular Psychiatry, 1–8. doi:10.1038/mp.2017.154 (2017).

30. Demontis D et al. Discovery of the first genome-wide significant risk loci for attention deficit/hyperactivity disorder. Nature Genetics 51, 63–75. doi:10.1038/s41588-018-0269-7 (2019).

31. Wray NR et al. Genome-wide association analyses identify 44 risk variants and refine the genetic architecture of major depression. Nature Genetics 50, 668–681. doi:10.1038/s41588-018-0090-3 (2018).

32. The Autism Spectrum Disorders Working Group of The Psychiatric Genomics Consortium. Metaanalysis of GWAS of over 16,000 individuals with autism spectrum disorder highlights a novel locus at 10q24.32 and a significant overlap with schizophrenia. Molecular autism 8, 21. doi:10.1186/s13229-017-0137-9 (2017).

33. Otowa T et al. Meta-analysis of genome-wide association studies of anxiety disorders. Molecular Psychiatry 21, 1391–1399. doi:10.1038/mp.2015.197 (2016).

34. Kanai M et al. Genetic analysis of quantitative traits in the Japanese population links cell types to complex human diseases. Nature genetics 50, 390–400. doi:10.1038/s41588-018-0047-6 (2018).

35. Choi SW, O’Reilly PF. PRSice-2: Polygenic Risk Score software for biobank-scale data. GigaScience 8, 1–6. doi:10.1093/gigascience/giz082 (2019).

36. Watanabe K, Taskesen E, Van Bochoven A, Posthuma D. Functional mapping and annotation of genetic associations with FUMA. Nature Communications 8. doi:10.1038/s41467-017-01261-5 (2017).

37. The GTEx Consortium. The Genotype-Tissue Expression (GTEx) project. Nature genetics 45, 580–5. doi:10.1038/ng.2653 (2013).

38. GTEx Consortium. The GTEx Consortium atlas of genetic regulatory effects across human tissues. Science (New York, N.Y.) 369, 1318–1330. doi:10.1126/science.aaz1776 (2020).

39. Wang Y et al. The 3D Genome Browser: A web-based browser for visualizing 3D genome organization and long-range chromatin interactions. Genome Biology 19, 151. doi:10.1186/s13059-018-1519-9 (2018).

40. Jung I et al. A compendium of promoter-centered long-range chromatin interactions in the human genome. Nature Genetics 51, 1442–1449. doi:10.1038/s41588-019-0494-8 (2019).

41. Purcell S et al. PLINK: A Tool Set for Whole-Genome Association and Population-Based Linkage Analyses. The American Journal of Human Genetics 81, 559–575. doi:10.1086/519795 (2007).

42. Chang CC, Chow CC, Tellier LC, Vattikuti S, Purcell SM, Lee JJ. Second-generation PLINK: rising to the challenge of larger and richer datasets. GigaScience 4, 7. doi:10.1186/s13742-015-0047-8 (2015).

43. De Leeuw C et al. Involvement of astrocyte metabolic coupling in Tourette syndrome pathogenesis. European Journal of Human Genetics 23, 1519–1522. doi:10.1038/ejhg.2015.22 (2015).

44. Bycroft C et al. The UK Biobank resource with deep phenotyping and genomic data. Nature 562, 203–209. doi:10.1038/s41586-018-0579-z (2018).

45. Euesden J, Lewis CM, O’Reilly PF. PRSice: Polygenic Risk Score software. Bioinformatics 31, 1466–1468. doi:10.1093/bioinformatics/btu848 (2015).

46. Anttila V et al. Analysis of shared heritability in common disorders of the brain. Science 360, eaap8757. doi:10.1126/science.aap8757 (2018).

47. Tylee DS et al. Genetic correlations among psychiatric and immune-related phenotypes based on genome-wide association data. American Journal of Medical Genetics Part B: Neuropsychiatric Genetics 177, 641–657. doi:10.1002/ajmg.b.32652 (2018).

48. Mufford M et al. Concordance of genetic variation that increases risk for Tourette Syndrome and that influences its underlying neurocircuitry. Translational Psychiatry 9, 120. doi:10.1038/s41398-019-0452-3 (2019).

49. Goodart SA, Butler MG, Overhauser J. Familial double pericentric inversion of chromosome 5 with some features of cri-du-chat syndrome. Human Genetics 97, 802–807. doi:10.1007/BF02346193 (1996).

50. Malan V et al. Molecular characterisation of a prenatally diagnosed 5q15q21.3 deletion and review of the literature 2006. doi:10.1002/pd.1386.

51. Brown KK, Alkuraya FS, Matos M, Robertson RL, Kimonis VE, Morton CC. NR2F1 deletion in a patient with a de novo paracentric inversion, inv(5)(q15q33.2), and syndromic deafness. American Journal of Medical Genetics, Part A 149, 931–938. doi:10.1002/ajmg.a.32764 (2009).

52. Al-Kateb H, Shimony JS, Vineyard M, Manwaring L, Kulkarni S, Shinawi M. NR2F1 haploinsufficiency is associated with optic atrophy, dysmorphism and global developmental delay 2013. doi:10.1002/ajmg.a.35650.

53. Smith C, Bolton A, Nguyen G. Genomic and Epigenomic Instability, Fragile Sites, Schizophrenia and Autism. Current Genomics 11, 447–469. doi:10.2174/138920210793176001 (2010).

54. Ang CE et al. The novel lncRNA lnc-NR2F1 is pro-neurogenic and mutated in human neurodevelopmental disorders. eLife 8 (eds West AE, Bronner ME) e41770. doi:10.7554/eLife.41770 (2019).

55. Bertacchi M et al. Mouse *Nr2f1* haploinsufficiency unveils new pathological mechanisms of a human optic atrophy syndrome. EMBO Molecular Medicine 11. doi:10.15252/emmm.201910291 (2019).

56. Zhang K et al. Imbalance of Excitatory/Inhibitory Neuron Differentiation in Neurodevelopmental Disorders with an NR2F1 Point Mutation. Cell Reports 31, 107521. doi:10.1016/j.celrep.2020.03.085 (2020).

57. Del Pino I et al. COUP-TFI/Nr2f1 Orchestrates Intrinsic Neuronal Activity during Development of the Somatosensory Cortex. Cerebral Cortex 30, 5667–5685. doi:10.1093/cercor/bhaa137 (2020).

58. Manikandan M et al. NR2F1 mediated down-regulation of osteoblast differentiation was rescued by bone morphogenetic protein-2 (BMP-2) in human MSC. Differentiation 104, 36–41. doi:10.1016/j.diff.2018.10.003 (2018).

59. Rech ME et al. Phenotypic expansion of Bosch-Boonstra-Schaaf optic atrophy syndrome and further evidence for genotype-phenotype correlations. American journal of medical genetics. Part A 182, 1426–1437. doi:10.1002/AJMG.A.61580 (2020).

60. Perets R et al. Genome-Wide Analysis of Androgen Receptor Targets Reveals COUP-TF1 as a Novel Player in Human Prostate Cancer. PLoS ONE 7. doi:10.1371/journal.pone.0046467 (2012).

61. Sosa MS et al. NR2F1 controls tumour cell dormancy via SOX9and RARβ -driven quiescence programmes. Nature Communications 6, 6170. doi:10.1038/ncomms7170 (2015).

62. Tang K, Tsai SY, Tsai MJ. COUP-TFs and eye development 2015. doi:10.1016/j.bbagrm.2014.05.022.

63. Shibata H et al. COUP-TFI expression in human adrenocortical adenomas: Possible role in steroidogenesis. Journal of Clinical Endocrinology and Metabolism 83, 4520–4523. doi:10.1210/jcem.83.12.5470 (1998).

64. Vuckovic D et al. The Polygenic and Monogenic Basis of Blood Traits and Diseases. Cell 182, 1214–1231.e11. doi:10.1016/j.cell.2020.08.008 (2020).

65. Chen MH et al. Trans-ethnic and Ancestry-Specific Blood-Cell Genetics in 746,667 Individuals from 5 Global Populations. Cell 182, 1198–1213.e14. doi:10.1016/j.cell.2020.06.045 (2020).

66. Lee JJ et al. Gene discovery and polygenic prediction from a genome-wide association study of educational attainment in 1.1 million individuals. Nature genetics 50, 1112–1121. doi:10.1038/s41588-018-0147-3 (2018).

67. Liu M et al. Association studies of up to 1.2 million individuals yield new insights into the genetic etiology of tobacco and alcohol use 2019. doi:10.1038/s41588-018-0307-5.

68. Yin M et al. Transactivation of microRNA-383 by steroidogenic factor-1 promotes estradiol release from mouse ovarian granulosa cells by targeting RBMS1. Molecular Endocrinology 26, 1129–1143. doi:10.1210/me.2011-1341 (2012).

69. Azhar S, Dong D, Shen WJ, Hu Z, Kraemer FB. The role of miRNAs in regulating adrenal and gonadal steroidogenesis 2020. doi:10.1530/JME-19-0105.

70. Peterson BS et al. Steroidhormonesand CNS sexual dimorphisms modulate symptom expression in tourette’s syndrome 1992. doi:10.1016/0306-4530(92)90015-Y.

71. Martino D, Macerollo A, Leckman JF. Neuroendocrine aspects of tourette syndrome 1st ed., 239–279. doi:10.1016/B978-0-12-411546-0.00009-3 (Elsevier Inc., 2013).

72. Corbett BA, Mendoza SP, Baym CL, Bunge SA, Levine S. Examining cortisol rhythmicity and responsivity to stress in children with Tourette syndrome. Psychoneuroendocrinology 33, 810–820. doi:10.1016/j.psyneuen.2008.03.014 (2008).

73. Buse J, Kirschbaum C, Leckman JF, Münchau A, Roessner V. The Modulating Role of Stress in the Onset and Course of Tourette’s Syndrome: A Review. Behavior Modification 38, 184–216. doi:10.1177/0145445514522056 (2014).

