## Supplement for "Genome-wide Association Study identifies two novel loci for Gilles de la Tourette Syndrome"

Supplementary Table 1: Listing of the sources of the samples used in this study, and the country of origin.

| Source | Cases | Controls | Platform |
|---|---|---|---|
| 3-City Study | 0 | 500 | Human610-Quad |
| Ashkenazi-Jewish controls from Yeshiva University | 0 | 1227 | HumanOmniExpress-12 |
| This study | 1633 | 2934 | HumanOmniExpress-24 |
| | | | OmniExpressExome-8 |
| POPGEN study | 0 | 675 | HumanHap550 |
| Spanish controls | 0 | 235 | CustomNeurochipHumanCore-24 |
| Swedish cohort | 52 | 58 | OmniExpressExome-8 |
| 1958 Birth Cohort from WTCCC | 0 | 2929 | Human1.2M-Duo |

| Country | Samples | Platform |
|---|---|---|
| Albania | 42 | HumanOmniExpress-24, HumanOmni2.5M |
| Denmark | 1306 | HumanOmniExpress-12, HumanOmniExpress-24 |
| France | 628 | HumanOmniExpress-24, OmniExpressExome-8, Human610-Quad |
| Germany | 1093 | HumanOmniExpress-24, OmniExpressExome-8, HumanHap550 |
| Greece | 660 | HumanOmniExpress-24, OmniExpressExome-8, HumanOmni2.5M |
| Hungary | 530 | HumanOmniExpress-24, OmniExpressExome-8 |
| Israel | 1275 | HumanOmniExpress-12, HumanOmniExpress-24 |
| Italy | 323 | HumanOmniExpress-24, OmniExpressExome-8, HumanOmni2.5M |
| Netherlands | 90 | HumanOmniExpress-24 |
| Poland | 858 | HumanOmniExpress-24 |
| Spain | 320 | HumanOmniExpress-24, HumanOmni2.5M, CustomNeurochipHumanCore-24 |
| Sweden | 110 | OmniExpressExome-8 |
| Switzerland | 22 | HumanOmniExpress-24 |
| UnitedKingdom | 2986 | Human1.2M-Duo, HumaOmniExpress-24 |

Supplementary Table 2: ANOVA statistics by EIGENSOFT on case/control status in the data. With three crosses we indicate the eigenvectors that are significantly associated with population structure. Four eigenvectors were identified as significant, Eigenvectors 1, 2, 4, and 5.

| **Eigenvector** | **P-value** | **Significance** |
|---|---|---|
| 1 | 1.45918E-11 | +++ |
| 2 | 4.57412E-13 | +++ |
| 3 | 0.536148 | |
| 4 | 8.18456E-12 | +++ |
| 5 | 0 | +++ |
| 6 | 0.00210966 | |
| 7 | 0.310167 | |
| 8 | 0.0762842 | |
| 9 | 0.176993 | |
| 10 | 0.975668 | |
| 11 | 0.608711 | |
| 12 | 0.268887 | |
| 13 | 0.334205 | |
| 14 | 0.918717 | |
| 15 | 0.0620558 | |
| 16 | 0.707506 | |
| 17 | 0.58127 | |
| 18 | 0.960909 | |
| 19 | 1 | |
| 20 | 0.0450066 | |

Supplementary Table 3: Replication of the gene sets identified as significant in previous publications. All gene sets attained significance. NSNP is the number of SNPs contained in the genes of the gene set. NSIG is the number of SNPs attaining nominal significance. EMP1 is the empirical p-value produced by the test.

| SET | NSNP | NSIG | EMP1 |
|---|---|---|---|
| Cell adhesion and transsynaptic signaling | 1939 | 369 | 0.00009999 |
| Lymphocytes | 623 | 122 | 0.00009999 |
| Astrocyte-neuron metabolic coupling | 147 | 20 | 0.0005999 |
| Ligand-gated Ion Channel Signaling | 432 | 81 | 0.0005999 |

Supplementary Table 4: Results from partitioned LDSC heritability analysis using the extended sets of the baseline model. Coefficient SE is the standard error of the coefficient. Coefficient P is the p-value. After Benjamini-Hochberg FDR correction, H3K4me1 and Conserved elements were identified as statistically significant.

| **Category** | **Prop.SNPs** | **Prop.$H^2$** | **Prop.$H^2$SE** | **Enrich.** | **Enrich.SE** | **Enrich.P** | **Coeff.** | **Coeff.SE** | **Coeff.Z** |
|---|---|---|---|---|---|---|---|---|---|
| H3K4me1_Trynka | 0.606 | 0.974 | 0.104 | 1.607 | 0.171 | 9.55E-04 | 9.40E-08 | 7.61E-08 | 1.235 |
| Conserved_LindbladToh | 0.330 | 0.678 | 0.120 | 2.052 | 0.363 | 3.85E-03 | 2.82E-08 | 4.10E-08 | 0.689 |
| H3K27ac_PGC2 | 0.335 | 0.668 | 0.133 | 1.993 | 0.397 | 9.38E-03 | 1.83E-07 | 1.55E-07 | 1.177 |
| DGF_ENCODE | 0.538 | 0.149 | 0.189 | 0.277 | 0.351 | 2.63E-02 | -2.10E-07 | 7.14E-08 | -2.940 |
| Enhancer_Andersson | 0.019 | 0.149 | 0.062 | 7.852 | 3.264 | 3.70E-02 | 4.96E-07 | 2.90E-07 | 1.706 |
| DHS_Trynka | 0.496 | 0.828 | 0.195 | 1.669 | 0.393 | 9.88E-02 | 1.93E-07 | 9.74E-08 | 1.984 |
| FetalDHS_Trynka | 0.283 | 0.528 | 0.185 | 1.865 | 0.654 | 1.89E-01 | 3.64E-08 | 9.66E-08 | 0.377 |
| Enhancer_Hoffman | 0.090 | -0.035 | 0.112 | -0.384 | 1.246 | 2.55E-01 | 1.58E-09 | 1.92E-07 | 0.008 |
| Intron_UCSC | 0.397 | 0.463 | 0.063 | 1.167 | 0.159 | 2.79E-01 | -4.22E-07 | 4.83E-07 | -0.873 |
| SuperEnhancer_Hnisz | 0.170 | 0.237 | 0.059 | 1.391 | 0.348 | 2.84E-01 | -4.60E-07 | 7.80E-07 | -0.590 |
| TFBS_ENCODE | 0.341 | 0.163 | 0.189 | 0.477 | 0.555 | 3.34E-01 | -5.39E-08 | 9.02E-08 | -0.598 |
| H3K9ac_Trynka | 0.230 | 0.358 | 0.136 | 1.558 | 0.592 | 3.40E-01 | 2.46E-08 | 1.13E-07 | 0.217 |
| H3K27ac_Hnisz | 0.420 | 0.501 | 0.103 | 1.191 | 0.245 | 4.41E-01 | 8.31E-08 | 2.06E-07 | 0.404 |
| TSS_Hoffman | 0.034 | -0.012 | 0.075 | -0.352 | 2.188 | 5.32E-01 | 8.64E-09 | 2.89E-07 | 0.030 |
| UTR_5_UCSC | 0.027 | -0.016 | 0.069 | -0.584 | 2.561 | 5.36E-01 | 7.16E-08 | 2.19E-07 | 0.327 |
| H3K4me3_Trynka | 0.255 | 0.313 | 0.156 | 1.228 | 0.611 | 7.09E-01 | 2.13E-08 | 1.04E-07 | 0.206 |
| Transcribed_Hoffman | 0.762 | 0.719 | 0.118 | 0.943 | 0.155 | 7.14E-01 | -2.06E-08 | 4.90E-08 | -0.422 |
| CTCF_Hoffman | 0.071 | 0.112 | 0.127 | 1.578 | 1.787 | 7.46E-01 | 1.61E-07 | 1.95E-07 | 0.827 |
| UTR_3_UCSC | 0.026 | 0.038 | 0.050 | 1.442 | 1.904 | 8.17E-01 | -2.03E-07 | 1.87E-07 | -1.089 |
| Coding_UCSC | 0.064 | 0.078 | 0.073 | 1.230 | 1.154 | 8.42E-01 | -5.24E-08 | 1.17E-07 | -0.448 |
| WeakEnhancer_Hoffman | 0.089 | 0.067 | 0.119 | 0.755 | 1.346 | 8.55E-01 | 1.62E-08 | 1.31E-07 | 0.124 |
| Promoter_UCSC | 0.038 | 0.046 | 0.055 | 1.217 | 1.451 | 8.81E-01 | -4.10E-09 | 5.06E-07 | -0.008 |
| Repressed_Hoffman | 0.719 | 0.730 | 0.081 | 1.016 | 0.113 | 8.85E-01 | -2.73E-08 | 5.79E-08 | -0.471 |
| PromoterFlanking_Hoffman | 0.033 | 0.032 | 0.092 | 0.964 | 2.787 | 9.90E-01 | 3.95E-08 | 2.49E-07 | 0.159 |

Supplementary Table 5: Statistically significant results from partitioned LDSC heritability analysis using the annotation based on Roadmap epigenomic modification data. Coefficient SE is the standard error of the coefficient. Coefficient P is the p-value. BENJ is Boolean descriptor of passing Benjamini-Hochberg FDR threshold, BONF is Boolean descriptor of passing Bonferroni threshold. Total number of annotations contained in the dataset 396. Correction using Benjamini-Hochberg identified 41 statistically significant associations, with 30 being in brain tissues and 9 in blood-related cells.

| Name | Coefficient | Coefficient SE | Coefficient P | BENJ | BONF |
|---|---|---|---|---|---|
| Brain Inferior Temporal Lobe H3K9ac | 9.89E-07 | 2.29E-07 | 8.07E-06 | YES | YES |
| Brain Angular Gyrus H3K27ac | 5.00E-07 | 1.19E-07 | 1.22E-05 | YES | YES |
| Brain Angular Gyrus H3K4me1 | 5.76E-07 | 1.37E-07 | 1.33E-05 | YES | YES |
| Brain Inferior Temporal Lobe H3K27ac | 4.01E-07 | 9.56E-08 | 1.35E-05 | YES | YES |
| Brain Dorsolateral Prefrontal Cortex H3K27ac | 5.74E-07 | 1.37E-07 | 1.38E-05 | YES | YES |
| Brain Dorsolateral Prefrontal Cortex H3K9ac | 1.27E-06 | 3.15E-07 | 2.79E-05 | YES | YES |
| Brain Anterior Caudate H3K9ac | 8.47E-07 | 2.18E-07 | 5.03E-05 | YES | YES |
| Brain Dorsolateral Prefrontal Cortex H3K4me1 | 6.07E-07 | 1.58E-07 | 5.82E-05 | YES | YES |
| Brain Angular Gyrus H3K9ac | 1.02E-06 | 2.69E-07 | 7.47E-05 | YES | YES |
| Brain Inferior Temporal Lobe H3K4me1 | 6.05E-07 | 1.62E-07 | 9.71E-05 | YES | YES |
| Brain Cingulate Gyrus H3K4me1 | 4.74E-07 | 1.28E-07 | 1.05E-04 | YES | YES |
| Brain Cingulate Gyrus H3K27ac | 3.93E-07 | 1.09E-07 | 1.47E-04 | YES | NO |
| Brain Substantia Nigra H3K4me1 | 4.27E-07 | 1.18E-07 | 1.48E-04 | YES | NO |
| Fetal Brain Female H3K4me3 | 1.08E-06 | 3.04E-07 | 1.99E-04 | YES | NO |
| Brain Anterior Caudate H3K4me1 | 4.69E-07 | 1.35E-07 | 2.66E-04 | YES | NO |
| Brain Hippocampus Middle H3K27ac | 3.50E-07 | 1.02E-07 | 2.86E-04 | YES | NO |
| Brain Anterior Caudate H3K27ac | 3.61E-07 | 1.05E-07 | 3.05E-04 | YES | NO |
| Brain Hippocampus Middle H3K4me1 | 3.32E-07 | 9.72E-08 | 3.17E-04 | YES | NO |
| Brain Anterior Caudate H3K4me3 | 9.90E-07 | 2.93E-07 | 3.56E-04 | YES | NO |
| Brain Cingulate Gyrus H3K9ac | 9.75E-07 | 2.98E-07 | 5.33E-04 | YES | NO |
| Brain Substantia Nigra H3K27ac | 3.10E-07 | 9.93E-08 | 8.88E-04 | YES | NO |
| Brain Germinal Matrix H3K4me3 | 1.00E-06 | 3.24E-07 | 9.64E-04 | YES | NO |
| Brain Cingulate Gyrus H3K4me3 | 9.02E-07 | 2.92E-07 | 1.00E-03 | YES | NO |
| Brain Inferior Temporal Lobe H3K4me3 | 1.02E-06 | 3.30E-07 | 1.02E-03 | YES | NO |
| Brain Substantia Nigra H3K9ac | 8.86E-07 | 2.88E-07 | 1.04E-03 | YES | NO |
| Primary hematopoietic stem cells short term culture H3K4me3 | 9.86E-07 | 3.23E-07 | 1.14E-03 | YES | NO |
| Primary Natural Killer cells from peripheral blood H3K36me3 | 2.26E-07 | 7.62E-08 | 1.53E-03 | YES | NO |
| Brain Substantia Nigra H3K4me3 | 9.36E-07 | 3.21E-07 | 1.77E-03 | YES | NO |
| Ganglion Eminence derived primary cultured neurospheres H3K4me3 | 9.31E-07 | 3.24E-07 | 2.03E-03 | YES | NO |
| Primary hematopoietic stem cells G-CSF-mobilized Male H3K36me3 | 2.03E-07 | 7.15E-08 | 2.24E-03 | YES | NO |
| Brain Hippocampus Middle H3K4me3 | 7.27E-07 | 2.62E-07 | 2.72E-03 | YES | NO |
| Skeletal Muscle Female H3K9ac | 6.46E-07 | 2.36E-07 | 3.11E-03 | YES | NO |
| Brain Angular Gyrus H3K4me3 | 9.00E-07 | 3.29E-07 | 3.16E-03 | YES | NO |
| Primary hematopoietic stem cells H3K36me3 | 5.15E-07 | 1.89E-07 | 3.23E-03 | YES | NO |
| Primary T killer naive cells from peripheral blood H3K9ac | 1.25E-06 | 4.62E-07 | 3.52E-03 | YES | NO |
| Primary T cells from peripheral blood H3K36me3 | 1.69E-07 | 6.35E-08 | 3.81E-03 | YES | NO |
| Fetal Brain Female DNase | 7.22E-07 | 2.72E-07 | 3.96E-03 | YES | NO |
| Primary T killer naive cells from peripheral blood H3K36me3 | 3.50E-07 | 1.32E-07 | 4.09E-03 | YES | NO |
| Fetal Brain Male DNase | 6.62E-07 | 2.55E-07 | 4.72E-03 | YES | NO |
| Primary T helper cells PMA-I stimulated H3K36me3 | 1.60E-07 | 6.19E-08 | 4.80E-03 | YES | NO |
| Primary B cells from peripheral blood H3K36me3 | 1.72E-07 | 6.64E-08 | 4.81E-03 | YES | NO |

Supplementary Table 6: Results of the gene-based analysis in FUMA. None of the genes attained significance, with RANGAP1 being the top hit and approaching the significance threshold. Total number of genes examined 5,764. NSNPs is the number of SNPs associated with the respective gene. NPARAM is the number of relevant parameters used for the model. ZSTAT is the gene-test association statistic. P is the p-value.

| SYMBOL | CHR | START | STOP | NSNPS | NPARAM | ZSTAT | P |
|---|---|---|---|---|---|---|---|
| RANGAP1 | 22 | 41621615 | 41702255 | 39 | 5 | 4.5024 | 3.3591E-06 |
| PCDH7 | 4 | 30702037 | 31168422 | 416 | 28 | 4.2627 | 0.0000101 |
| AL035681.1 | 22 | 41665388 | 41705686 | 20 | 3 | 4.1834 | 0.000014362 |
| EFNA5 | 5 | 106692590 | 107026596 | 141 | 31 | 4.0205 | 0.000029036 |
| NCKIPSD | 3 | 48681364 | 48743797 | 23 | 4 | 3.9244 | 0.000043464 |
| CDKN1A | 6 | 36624305 | 36675116 | 83 | 10 | 3.8097 | 0.000069581 |
| PAM | 5 | 102069685 | 102386809 | 409 | 13 | 3.7867 | 0.000076322 |
| IP6K2 | 3 | 48705436 | 48797786 | 57 | 5 | 3.725 | 0.000097662 |
| PEX2 | 8 | 77872494 | 77933280 | 62 | 7 | 3.7154 | 0.00010142 |
| DCAF12 | 9 | 34066385 | 34147397 | 45 | 5 | 3.6834 | 0.00011506 |
| SRSF7 | 2 | 38950741 | 38998636 | 19 | 3 | 3.648 | 0.00013217 |
| KLHL5 | 4 | 39026659 | 39148477 | 187 | 7 | 3.551 | 0.00019186 |
| C2orf49 | 2 | 105933816 | 105985668 | 77 | 9 | 3.5387 | 0.00020107 |
| PFKM | 12 | 48478922 | 48560187 | 121 | 10 | 3.5377 | 0.00020182 |
| SLC10A4 | 4 | 48465360 | 48511213 | 45 | 5 | 3.5273 | 0.00020988 |
| MYT1L | 2 | 1772885 | 2355032 | 225 | 35 | 3.4759 | 0.00025461 |
| FBXL17 | 5 | 107174736 | 107737799 | 483 | 25 | 3.4598 | 0.00027025 |
| SENP1 | 12 | 48416681 | 48520091 | 184 | 12 | 3.4555 | 0.00027467 |
| ZAR1 | 4 | 48472269 | 48516406 | 42 | 5 | 3.4301 | 0.00030167 |
| EMB | 5 | 49672026 | 49759082 | 6 | 1 | 3.4289 | 0.000303 |
| EP300 | 22 | 41467790 | 41596081 | 104 | 6 | 3.4186 | 0.00031467 |
| TMCC2 | 1 | 205177304 | 205262471 | 56 | 6 | 3.3548 | 0.00039708 |
| ZNF382 | 19 | 37075719 | 37139499 | 74 | 6 | 3.3325 | 0.00043037 |
| MPO | 17 | 56327217 | 56378296 | 6 | 2 | 3.3306 | 0.00043336 |
| MCHR2 | 6 | 100347786 | 100462123 | 40 | 8 | 3.3216 | 0.00044747 |
| PPP1R3A | 7 | 113496832 | 113735975 | 113 | 3 | 3.3203 | 0.00044959 |
| DHRS11 | 17 | 34928228 | 34977235 | 7 | 2 | 3.2964 | 0.00048969 |
| MRM1 | 17 | 34938001 | 34985407 | 3 | 1 | 3.2954 | 0.00049132 |
| ZNF461 | 19 | 37108094 | 37177755 | 92 | 8 | 3.2544 | 0.00056807 |
| RP11-503N18.3 | 4 | 2431700 | 2484668 | 7 | 2 | 3.2387 | 0.00060036 |
| UBAP2 | 9 | 33901691 | 34068947 | 56 | 5 | 3.2239 | 0.00063232 |
| ARIH2OS | 3 | 48935221 | 48976818 | 27 | 3 | 3.2228 | 0.00063477 |
| RASGEF1A | 10 | 43669983 | 43782367 | 102 | 8 | 3.2197 | 0.00064153 |
| HOXB4 | 17 | 46632875 | 46677473 | 33 | 7 | 3.2098 | 0.00066407 |
| DNAJC22 | 12 | 49720700 | 49771309 | 8 | 1 | 3.2005 | 0.00068588 |
| PTPRU | 1 | 29543028 | 29673325 | 53 | 10 | 3.1858 | 0.00072183 |
| PRKAR2A | 3 | 48762030 | 48905279 | 63 | 4 | 3.1802 | 0.00073584 |
| ZC3H7B | 22 | 41677526 | 41776151 | 28 | 3 | 3.1796 | 0.00073748 |
| CADM2 | 3 | 84988132 | 86143579 | 1494 | 37 | 3.1738 | 0.00075229 |

Supplementary Table 7: Statistically significant results from the PRS analysis on the UK-Biobank MRI dataset using the summary statistics results of the meta-analysis. Trait is the coded ID in the UK Biobank, with its description. PRS coeff is the coefficient of the PRS, SD stands for the standard deviation, T-stat the T-test statistic, and Reg$R^2$ is the regression $r^2$ factor between PRS computed under each threshold and imaging trait of interest, carried out with age, sex and top 5 PCs as covariates. PRS-P is the p-value of the regression. P-threshold is the threshold for SNP allocation to the bin. In red we indicate the statistically significant signal from the analysis.

| Trait | PRS coeff. | SD | T-stat | PRS-P | Reg $R^2$ | P-Threshold | Description |
|---|---|---|---|---|---|---|---|
| 25018 | -7.57E-01 | 2.43E-01 | -3.12E+00 | 1.83E-03 | 1.32E-01 | 1E-5 | Volume of pallidum (right) |
| 25020 | -6.70E-01 | 2.44E-01 | -2.75E+00 | 6.01E-03 | 1.27E-01 | 1E-5 | Volume of hippocampus (right) |
| 25013 | -6.33E-01 | 2.45E-01 | -2.58E+00 | 9.77E-03 | 1.19E-01 | 1E-5 | Volume of caudate (left) |
| 25014 | -5.26E-01 | 2.46E-01 | -2.14E+00 | 3.25E-02 | 1.11E-01 | 1E-5 | Volume of caudate (right) |
| 25016 | -4.60E-01 | 2.24E-01 | 6.00E+00 | 3.98E-02 | 2.64E-01 | 1E-5 | Volume of putamen (right) |
| 25017 | -4.84E-01 | 2.44E-01 | -1.98E+00 | 4.73E-02 | 1.23E-01 | 1E-5 | Volume of pallidum (left) |
| 25021 | -4.92E-01 | 2.51E-01 | -1.96E+00 | 4.97E-02 | 7.57E-02 | 1E-5 | Volume of amygdala (left) |
| 25016 | -7.50E+00 | 1.82E+00 | -4.13E+00 | 3.70E-05 | 2.64E-01 | 1E-3 | Volume of putamen (right) |
| 25015 | -7.01E+00 | 1.83E+00 | -3.84E+00 | 1.25E-04 | 2.57E-01 | 1E-3 | Volume of putamen (left) |
| 25013 | -5.88E+00 | 1.99E+00 | -2.95E+00 | 3.15E-03 | 1.19E-01 | 1E-3 | Volume of caudate (left) |
| 25014 | -4.69E+00 | 2.00E+00 | -2.35E+00 | 1.89E-02 | 1.11E-01 | 1E-3 | Volume of caudate (right) |
| 25018 | -3.97E+00 | 1.97E+00 | 1.00E+00 | 4.47E-02 | 1.32E-01 | 1E-3 | Volume of pallidum (right) |

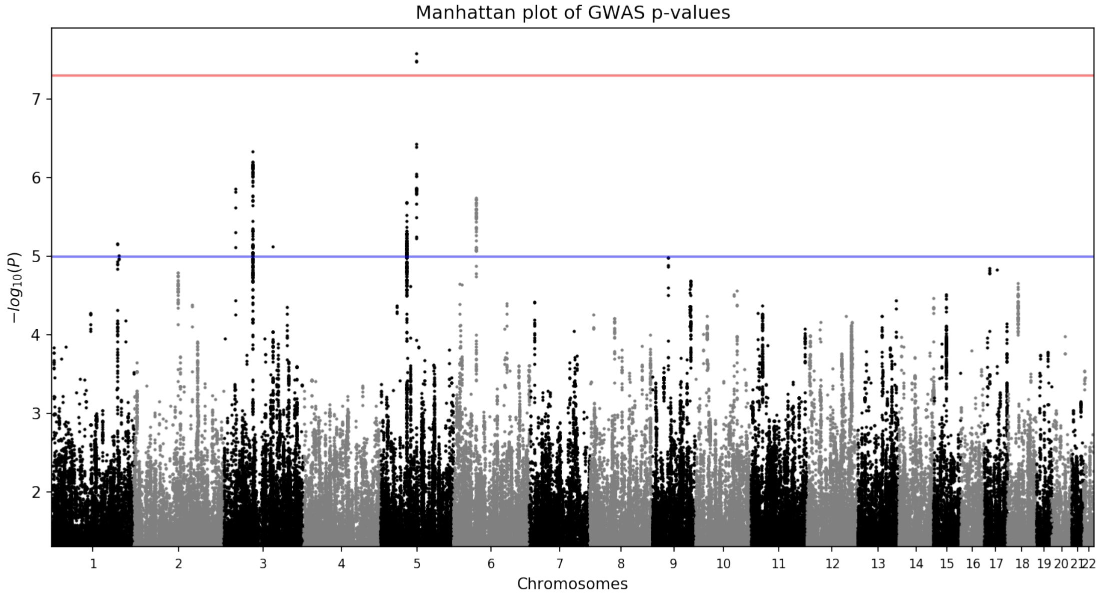

Supplementary Figure 1: The Manhattan plot for the TS-EUROTRAIN GWAS (1,438 cases and 4,356 controls on 2,949,675 variants). The -log10(p) values for the association tests (two-tailed) are shown on the y axis and the chromosomes are ordered on the x axis. One genetic locus on chromosome 5 surpassed the genome-wide significance threshold (p<5e-8 ; indicated by the red line). One genetic locus surpassed the array-wide significance threshold (p<1e-7). Gray and black differentiate adjacent chromosomes.

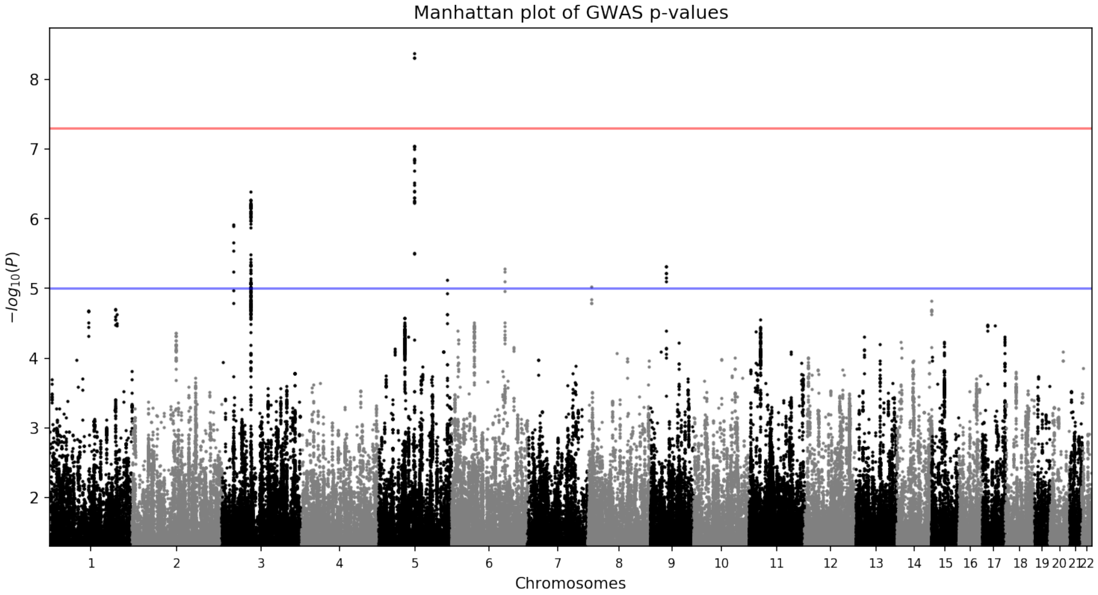

Supplementary Figure 2: The Manhattan plot for the the recomputed GWAS after excluding the overlapping samples between TS-EUROTRAIN and TSGWAS2 (1,314 cases and 4,077 controls on 2,949,675 variants). The -log10(p) values for the association tests (two-tailed) are shown on the y axis and the chromosomes are ordered on the x axis. One genetic locus on chromosome 5 surpassed the genome-wide significance threshold (p<5e-8 ; indicated by the red line). One genetic locus surpassed the array-wide significance threshold (p<1e-7). Gray and black differentiate adjacent chromosomes.

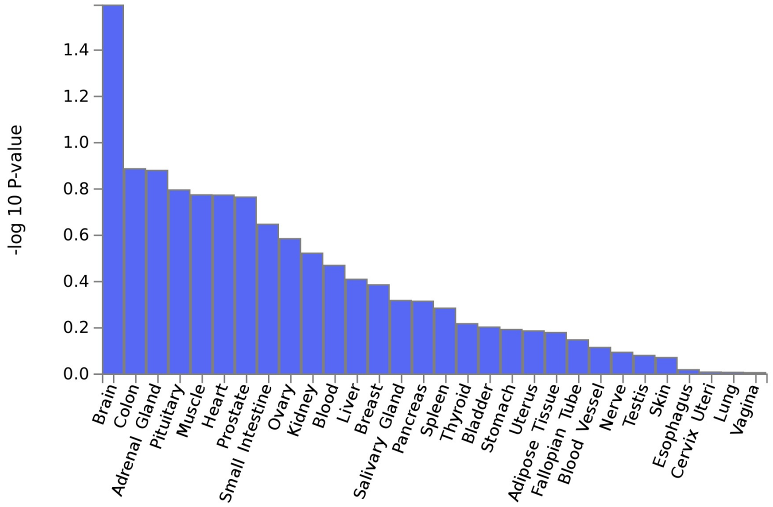

(a) Tissue enrichment for 30 tissues

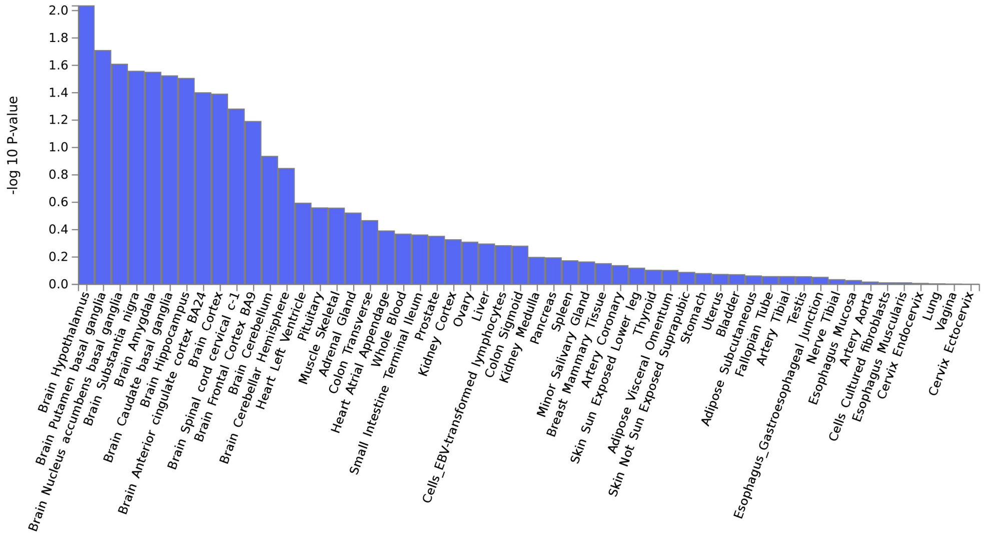

(b) Tissue enrichment for 53 tissues

Supplementary Figure 3: MAGMA tissue expression analysis on the TS-EUROTRAIN GWAS. a) Using 30 tissue samples from GTEx enrichment is indicated in the Brain, followed by the Colon, the Adrenal gland and the Pituitary. b) Using 53 tissue samples from GTEx enrichment is indicated in various brain tissues, with the top signals on Hypothalamus, Putamen basal ganglia, and Nucleus accumbens basal ganglia.

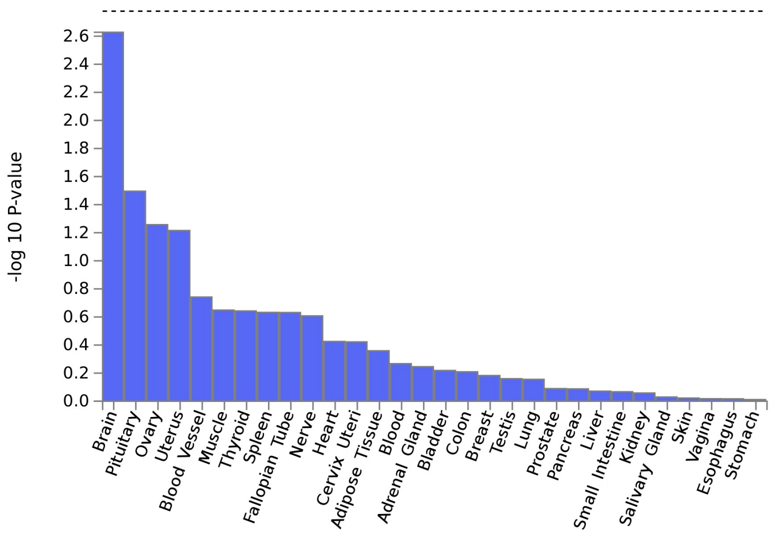

(a) Tissue enrichment for 30 tissues

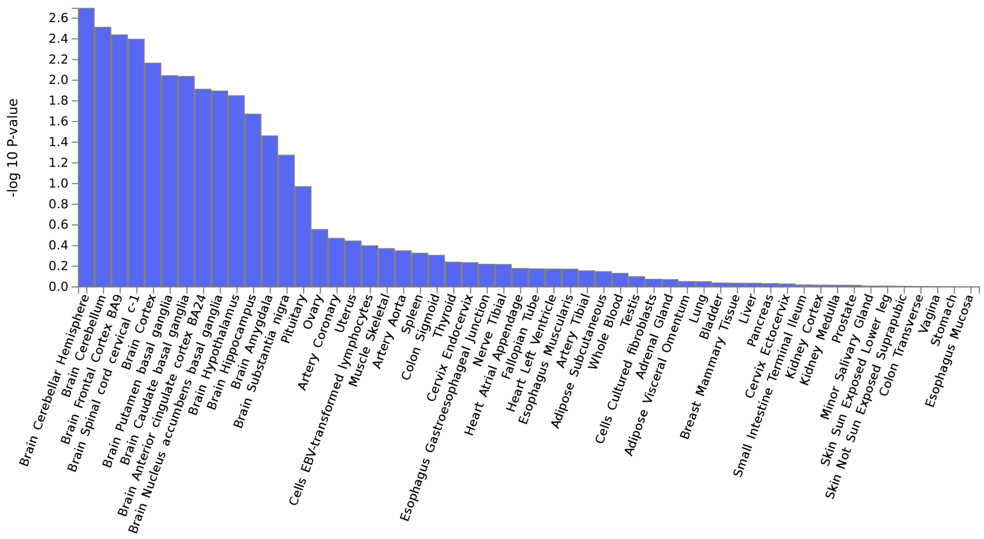

(b) Tissue enrichment for 53 tissues

Supplementary Figure 4: MAGMA tissue expression analysis on the meta-analysis. a) Using 30 tissue samples from GTEx, stronger enrichment can be discerned in the Brain, followed by the Pituitary and the Ovary. b) Using 53 tissue samples from GTEx stronger enrichment can be discerned for various brain tissues, with the top signals on Cerebellar Hemisphere, Cerebellum, Frontal Cortex BA9.

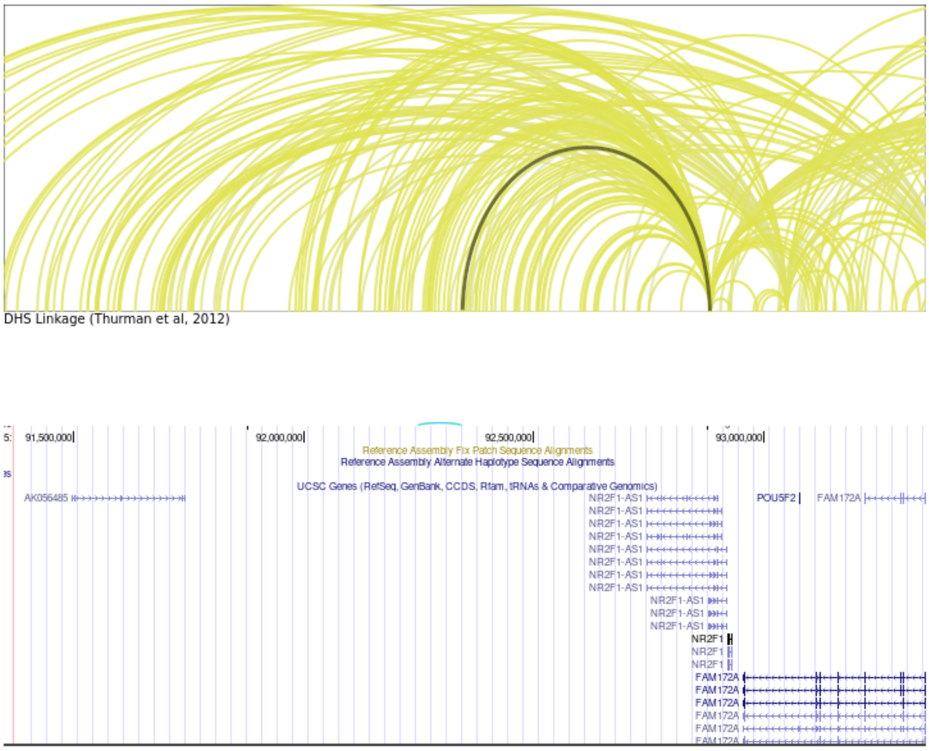

(a) Plot of the interactions identified by Capture Hi-C in the region around rs2453763. Highlighted is the interaction of rs2453763 with *NR2F1*.

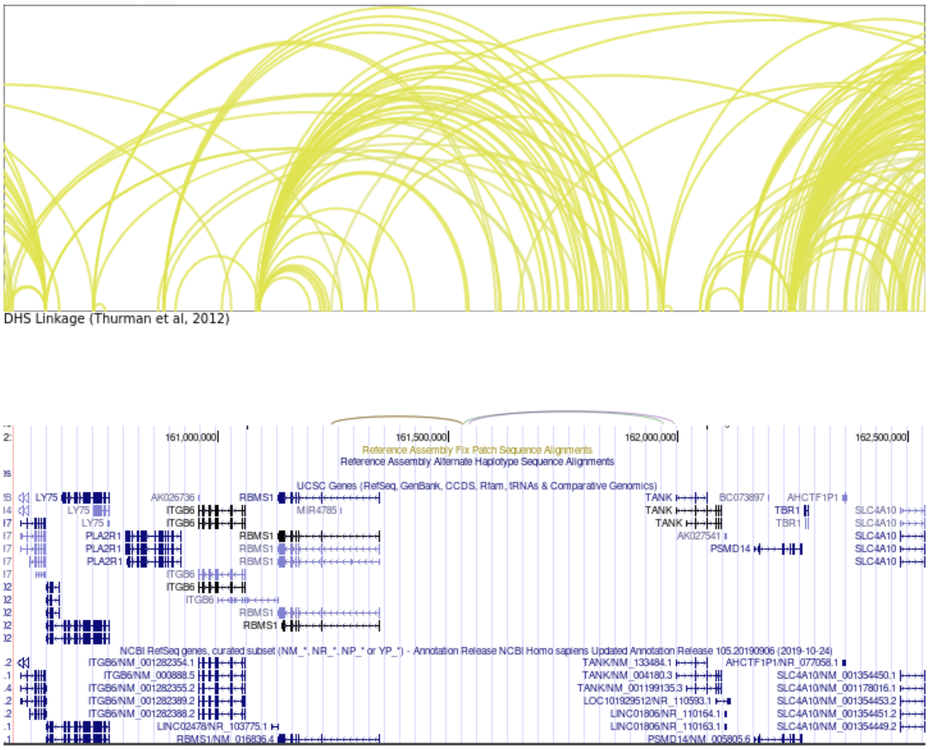

(b) Plot of the interactions identified by Capture Hi-C in the region around rs10209244.

Supplementary Figure 5: Capture Hi-C analysis for the top two variants of the meta-analysis.
